# Post-acute symptoms, new onset diagnoses and health problems 6 to 12 months after SARS-CoV-2 infection: a nationwide questionnaire study in the adult Danish population

**DOI:** 10.1101/2022.02.27.22271328

**Authors:** Anna Irene Vedel Sørensen, Lampros Spiliopoulos, Peter Bager, Nete Munk Nielsen, Jørgen Vinsløv Hansen, Anders Koch, Inger Kristine Meder, Steen Ethelberg, Anders Hviid

## Abstract

**Background:** A considerable number of individuals infected with SARS-CoV-2 continue to experience symptoms after the acute phase. More information on duration and prevalence of these symptoms in non-hospitalized populations is needed.

**Methods:** We conducted a nationwide cross-sectional study including 152 880 individuals aged 15-years or older, consisting of RT-PCR confirmed SARS-CoV-2 cases between September 2020-April 2021 (N=61 002) and a corresponding test-negative control group (N=91 878). Data were collected 6, 9 or 12 months after the test using web-based questionnaires. The questionnaire covered acute and post-acute symptoms, selected diagnoses, sick leave and general health, together with demographics and life style at baseline. Risk differences (RDs) between test-positives and -negatives were reported, adjusted for age, sex, single comorbidities, Charlson comorbidity score, obesity and healthcare-occupation.

**Findings:** Six to twelve months after the test date, the risks of 18 out of 21 physical symptoms were elevated among test-positives and one third (29.6%) of the test-positives experienced at least one physical post-acute symptom. The largest risk differences were observed for dysosmia (RD = 10.92%, 95%CI 10.68-11.21%), dysgeusia (RD=8.68%, 95%CI 8.43-8.93%), fatigue/exhaustion (RD=8.43%, 95%CI 8.14-8.74%), dyspnea (RD=4.87%, 95%CI 4.65-5.09%) and reduced strength in arms/legs (RD=4.68%, 95%CI 4.45-4.89%). More than half (53.1%) of test-positives reported at least one of the following conditions: concentration difficulties (RD=28.34%, 95%CI 27.34-28.78%), memory issues (RD=27.25%, 95%CI 26.80-27.71%), sleep problems (RD=17.27%, 95%CI 16.81-17.73%), mental (RD=32.58%, 95%CI 32.11-33.09%) or physical exhaustion (RD=40.45%, 95%CI 33.99-40.97%), compared to 11.5% of test-negatives. New diagnoses of anxiety (RD=1.15%, 95%CI 0.95-1.34%) or depression (RD=1.00%, 95%CI 0.81-1.19%) were also more common among test-positives.

**Interpretation:** At the population-level, where the majority of test-positives (96.0%) were not hospitalized during acute infection, a considerable proportion experience post-acute symptoms and sequelae 6-12 months after infection.

**Funding:** None

**Research in context:** *Evidence before the study:* To identify existing studies on the epidemiology and clinical nature of post-acute COVID-19 symptoms, we searched PubMed for articles published until January 4, 2022 using the search string (((SARS-CoV-2[Title/Abstract]) OR (COVID-19[Title/Abstract]) OR (coronavirus[Title/Abstract])) AND ((post-acute[Title/Abstract]) OR (“post acute”[Title/Abstract]) OR (“long haul*”[Title/Abstract]) OR (“long-term symptoms”[Title/Abstract]) OR (“long-term disease”[Title/Abstract]) OR (“long-term illness”[Title/Abstract]) OR (“persistent symptoms”[Title/Abstract]) OR (“persistent disease”[Title/Abstract]) OR (“persistent illness”[Title/Abstract]) OR (“prolonged symptoms”[Title/Abstract]) OR (“prolonged disease”[Title/Abstract]) OR (“prolonged illness”[Title/Abstract]))) OR (long-covid[Title/Abstract]) OR (“Post-COVID-19 syndrome”[Title/Abstract]) OR (“Post-COVID-19 condition”[Title/Abstract]) OR (“Post-COVID-19 symptoms”[Title/Abstract]). This resulted in 870 articles. When screening these, we focused on articles covering symptoms comprehensively or a broader area, e.g. mental health problems, rather than in-depth studies of symptoms within a single area, case stories or studies focusing on clinical management. A very wide range of post-acute symptoms originating from many different organ systems have been reported. This includes pulmonary, cardiovascular, hematologic, gastrointestinal, renal, endocrine, dermatologic, neurological and cognitive symptoms, as well as more general health problems, in particular fatigue. Based on two systematic reviews covering the period December 2019-March 2021, the majority of studies of persistent COVID-19 symptoms had until then been conducted among hospitalized patients and thus were not representative of the general population, where the majority will only have suffered mild or moderate disease. Since then some larger register-based studies as well as some smaller questionnaire- or interview-based studies have been conducted among non-hospitalized patients. Both types of studies offer advantages and disadvantages in relation to obtaining the full overview of long-term effects. Register-based studies are best suited for capturing more severe conditions confirmed by a trained physician and defined by diagnostic classification schemes, whereas questionnaires including self-reported symptoms are able to capture symptoms and health outcomes that do not easily confirm to disease diagnoses, but which are nevertheless critical to our understanding of the burden of post-acute symptoms. The number of sstudies of post-acute conditions among non-hospitalized patients with a follow-up time of more than 6 months is still limited. Some of the major remaining knowledge gaps regarding post-acute symptoms are: 1) What is the prevalence and variety of post-acute symptoms in the general population of infected persons, where the majority will only have suffered mild or moderate disease, 2) For how long do post-acute symptoms persist, and 3) Which subgroups of individuals, if any, are at higher risk of post-acute symptoms.

*Added value of this study:* The present nationwide questionnaire-study is based on a large, mainly adult study population (N=152 880), where all individuals in Denmark, who tested positive during the study period, were invited to participate along with comparable test-negative controls. Marked levels of post-acute symptoms and conditions were reported with changes in sense of smell and taste being the most frequently reported single physical symptoms. As many as half (53.1%) of the participants report having experienced general health problems in the form of either mental or physical exhaustion, sleep problems or cognitive problems, compared to 11.5% of control persons 6 to 12 months after the test. Our results suggest that a considerable proportion of the general population, who did not experience severe disease, are still affected 6 to 12 months after infection and that post-acute symptoms are more often experienced by females and middle-aged individuals.

*Implications of all available evidence:* Diverse post-acute symptoms following infection with SARS-CoV-2 occur frequently. Even up to 12 months after the onset of infection, a considerable proportion of individuals, who did not experience severe disease, continue to experience symptoms. Post-acute symptoms are generally more often reported by females than males, whereas the influence of age remains unclear.

## Introduction

A significant number of individuals infected with SARS-CoV-2 continue to experience symptoms after the acute phase of infection^1^. These symptoms have collectively been known under many different names including *long-COVID*, and has now been included in the WHO International Classification of Diseases under the name *post COVID-19 condition*^2^. Recently, the WHO established the following clinical case definition: “Post COVID-19 condition occurs in individuals with a history of probable or confirmed SARS-CoV-2 infection, usually 3 months from the onset of COVID-19 with symptoms that last for at least 2 months and cannot be explained by an alternative diagnosis”^3^.

The symptomatology of post COVID-19 condition is complex with possible involvement of multiple organ systems. A growing number of studies support that in addition to a wide range of unspecific physical symptoms, post-acute COVID symptoms may also comprise impaired cognition, mental health problems and chronic fatigue like conditions^4–6^. However, knowledge gaps remain regarding the prevalence, range and duration of these symptoms in the general population of infected and if subgroups particularly prone to post-acute symptoms exist.

To provide urgently needed insights into post COVID-19 condition, we conducted the, to the best of our knowledge, largest questionnaire-survey to date on long-COVID both globally and in the Danish population, the EFTER-COVID (Danish for AFTER-COVID) survey. In this report, we present results based on completed questionnaires from participants with a positive test for SARS-CoV-2 in the period September 2020 to April 2021 and corresponding test-negative controls.

The main objectives of this study were to: 1) Estimate the risk difference between SARS-CoV-2 test-positive and test-negative individuals for acute as well as post-acute symptoms 6-12 months after the test, 2) Evaluate the duration of post-acute symptoms, and 3) Explore the influence of age, sex and disease severity (hospitalisation) on post-acute symptoms.

## Methods

### Study design and population

In this nationwide cross-sectional survey, data on self-reported symptoms were collected using web-based questionnaires distributed via the national ‘e-Boks’ system, which is a platform offering electronic postal communication with public authorities (www.digst.dk). This system is used by 92% of all residents in Denmark aged 15-years and above.

In Denmark, free access to reverse transcription polymerase chain reaction (RT-PCR) tests for SARS-CoV-2 has been available for all adults since May 2020 independent of test indication^7^. Individuals invited to participate in the study were selected based on RT-PCR test results recorded in the national COVID-19 surveillance system at Statens Serum Institut, which captures the individual results of all RT-PCR tests performed (https://covid19.ssi.dk/). All individuals who tested positive during September 1, 2020 to April 2 2021, and who had an e-Boks account were invited to participate, along with controls in the form of individuals testing negative only during the same period. Controls were randomly selected using incidence density sampling on the test date with a ratio of 2:3 between test-positives and -negatives. This ratio was chosen to counteract a possible lower response rate among controls than cases. Individuals receiving more than one positive test result during the study period, where included based on the first result. Data were collected during August 1, 2021 to December 11, 2021, where participants received an invitation letter containing a link to the questionnaire 6, 9 or 12 months after their test date. Non-responders received a reminder 7-10 days after the invitation.

In order to minimize recall bias for acute symptoms, individuals with tests older than 12 months were not invited.

To avoid misclassification bias, controls, who reported having been found seropositive, were excluded. Participants were specifically asked to report any symptom that they might have experienced, no matter the reason for these, in order to avoid information bias from test-positives omitting non-COVID-19 symptoms.

### Data sources

Data were collected using questionnaires created in SurveyXact (www.surveyxact.dk), which could be completed using a PC, smartphone or tablet. The questionnaire included questions on height, weight, education, employment, smoking and drinking habits, physical activity, sick leave and symptoms in the time around the test date, defined as from one week before the test and until four weeks after. To evaluate post-acute COVID-19 symptoms, participants were asked about: 1) symptoms during the past 14 days, 2) selected health conditions diagnosed by a medical doctor before and after the test date, and 3) self-reported experiences of specific physical and mental health problems 6 months before and up to 6 to 12 months after testing. For the reported symptoms and health conditions, participants were also asked about whether they used to regularly experience these before the test. Test-negatives were asked about test indication and whether they suspected ever having had COVID-19. All questions in the questionnaire were mandatory, except height, weight and alcohol consumption. The questionnaire is available as supplementary material (Text S1).

In Denmark, individual-level data from different data sources can be linked using a unique identifier (the CPR-number) assigned in the Civil Registration System. Using the CPR-number, questionnaire data were supplemented with register-based information on age and sex, information on healthcare occupation from authorization data^8^ as well as information on comorbidities and hospitalizations from the Danish National Patient Register (DNPR)^9^. The DNPR contains information on in- and outpatient diagnoses coded using ICD-10, which made it possible to calculate Charlson Comorbidity Index scores. Hospitalizations were considered COVID-19 related, if the patient had received a positive test result within 14 days of admission, and had been registered with one of the ICD-codes: DB342, DB342A, DB972, DB972A, DB972B, DB972B1 or DB948A. Hospital-acquired infections with SARS-CoV-2 were not included.

### Statistical methods

The prevalence of conditions among test-positive and -negative individuals were compared using risk differences (RDs). Parametric g-computation^10^ on logistic regression was used to estimate RDs (with 95% confidence intervals) among the exposed with adjustment for completion time (6, 9 or 12 months), age, sex, obesity, comorbidities from the questionnaire, Charlson Comorbidity Index scores and healthcare occupation. Based on results of other studies^11–17^, these variables were considered potential confounders. Symptoms prior to the test were also adjusted for, and for diagnoses and health conditions, only new onsets, defined as conditions occurring after testing, but not in the 6 months leading up to, were taken into account.

The 95% confidence intervals were obtained through bootstrap random resampling with 1000 iterations. The R-packages “riskCommunicator”^18^ and “Forester”^19^ were used for modelling and generation of forest plots, respectively. We estimated RDs for the following conditions: 1) Acute symptoms in relation to the test date (only test-negatives, who reported symptoms compatible with COVID-19 as indication for testing, were included as test-negative in this analysis), 2) Post-acute symptoms during the 14 days prior to questionnaire completion 6, 9 or 12 months after the test, 3) New onset diagnoses of anxiety, chronic fatigue syndrome, depression, fibromyalgia and post-traumatic stress disorder (PTSD) confirmed by a medical doctor since the test, and 4) New onset of mental or physical exhaustion, concentration difficulties, memory issues or sleep problems since the test.

Main analyses were based on pooled data from 6, 9 or 12 months after tests and did not take time into account. Supplementary analyses were carried out at each of the three time points to examine if effects change time.

Charlson Comorbidity Index scores^20^ were calculated based on data for the past 5 years extracted from the DNPR^9^. Scores were included in analyses as 0, 1 or ≥2, since very few had scores above 2. In the questionnaire, participants were asked supplementary questions about relevant comorbidities commonly treated in primary care (table S1) and therefore unlikely to be listed in the DNPR. Presence of these comorbidities were included in analyses as dichotomous variables. Obesity was defined as BMI≥30 for individuals aged 18 years or above and for 15-17 year olds international cut-off points for obesity by sex and age were used^21^.

P-values in table 1, S1 and S2, were estimated using student’s t-test for continuous variables and Pearson’s Chi-squared test for categorical variables.

**Table 1:**
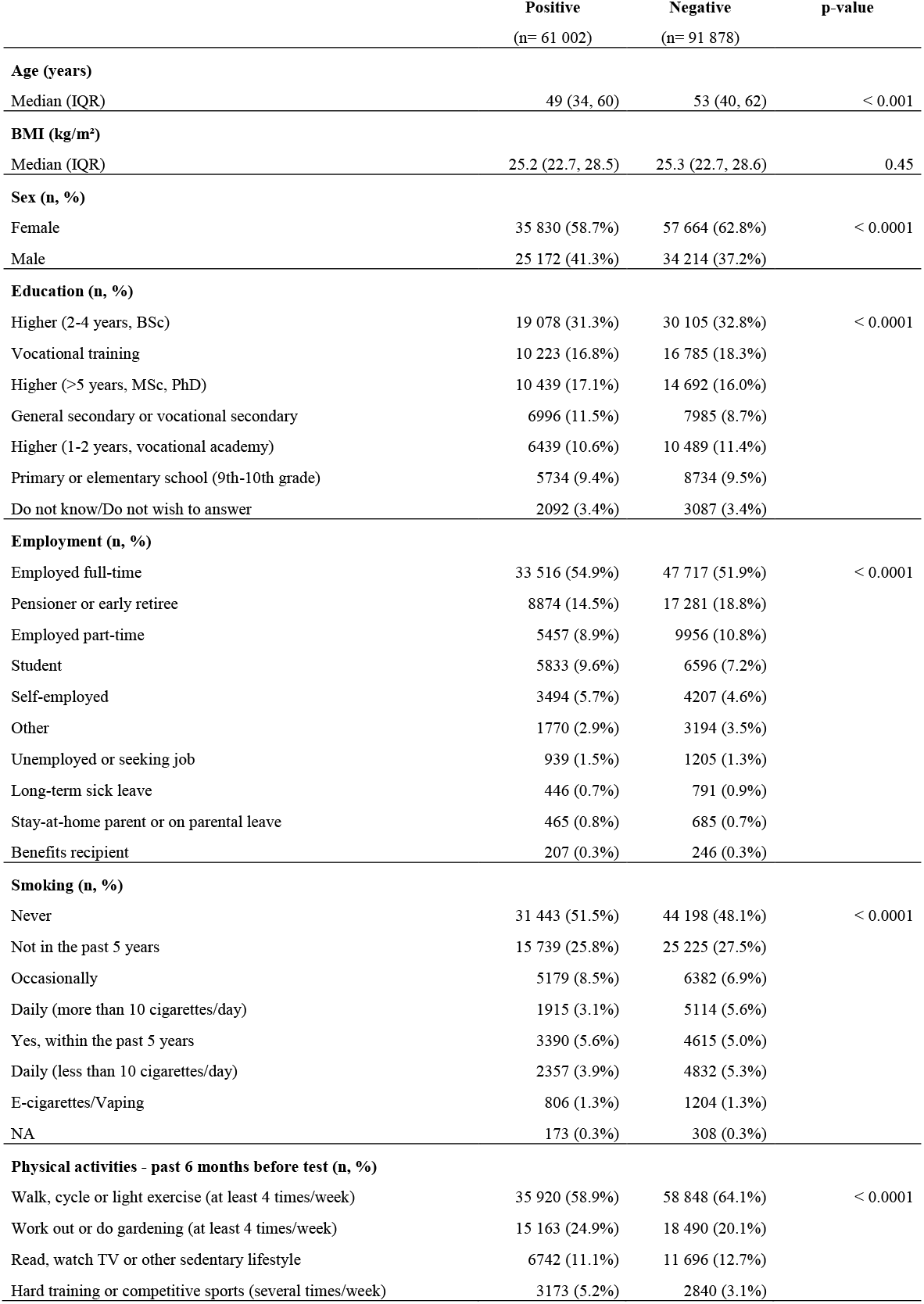
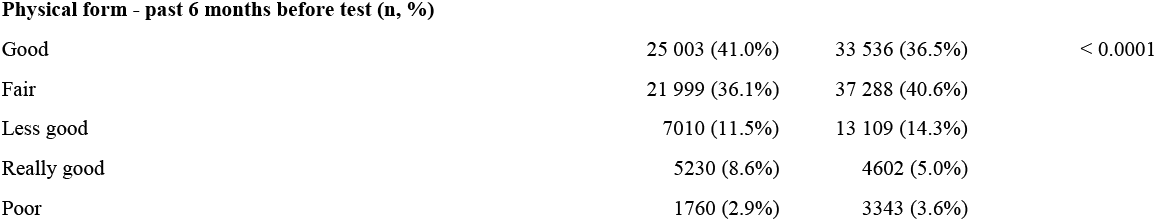
Characteristics of 152 880 survey participants tested for SARS-CoV-2 during September 1, 2020 – April 2, 2021.

|  | Positive<br>(n= 61 002) | Negative<br>(n= 91 878) | p-value |
| --- | --- | --- | --- |
| **Age (years)** |  |  |  |
| Median (IQR) | 49 (34, 60) | 53 (40, 62) | < 0.001 |
| **BMI (kg/m<sup>2</sup>)** |  |  |  |
| Median (IQR) | 25.2 (22.7, 28.5) | 25.3 (22.7, 28.6) | 0.45 |
| **Sex (n, %)** |  |  |  |
| Female | 35 830 (58.7%) | 57 664 (62.8%) | < 0.0001 |
| Male | 25 172 (41.3%) | 34 214 (37.2%) |  |
| **Education (n, %)** |  |  |  |
| Higher (2-4 years, BSc) | 19 078 (31.3%) | 30 105 (32.8%) | < 0.0001 |
| Vocational training | 10 223 (16.8%) | 16 785 (18.3%) |  |
| Higher (>5 years, MSc, PhD) | 10 439 (17.1%) | 14 692 (16.0%) |  |
| General secondary or vocational secondary | 6996 (11.5%) | 7985 (8.7%) |  |
| Higher (1-2 years, vocational academy) | 6439 (10.6%) | 10 489 (11.4%) |  |
| Primary or elementary school (9th-10th grade) | 5734 (9.4%) | 8734 (9.5%) |  |
| Do not know/Do not wish to answer | 2092 (3.4%) | 3087 (3.4%) |  |
| **Employment (n, %)** |  |  |  |
| Employed full-time | 33 516 (54.9%) | 47 717 (51.9%) | < 0.0001 |
| Pensioner or early retiree | 8874 (14.5%) | 17 281 (18.8%) |  |
| Employed part-time | 5457 (8.9%) | 9956 (10.8%) |  |
| Student | 5833 (9.6%) | 6596 (7.2%) |  |
| Self-employed | 3494 (5.7%) | 4207 (4.6%) |  |
| Other | 1770 (2.9%) | 3194 (3.5%) |  |
| Unemployed or seeking job | 939 (1.5%) | 1205 (1.3%) |  |
| Long-term sick leave | 446 (0.7%) | 791 (0.9%) |  |
| Stay-at-home parent or on parental leave | 465 (0.8%) | 685 (0.7%) |  |
| Benefits recipient | 207 (0.3%) | 246 (0.3%) |  |
| **Smoking (n, %)** |  |  |  |
| Never | 31 443 (51.5%) | 44 198 (48.1%) | < 0.0001 |
| Not in the past 5 years | 15 739 (25.8%) | 25 225 (27.5%) |  |
| Occasionally | 5179 (8.5%) | 6382 (6.9%) |  |
| Daily (more than 10 cigarettes/day) | 1915 (3.1%) | 5114 (5.6%) |  |
| Yes, within the past 5 years | 3390 (5.6%) | 4615 (5.0%) |  |
| Daily (less than 10 cigarettes/day) | 2357 (3.9%) | 4832 (5.3%) |  |
| E-cigarettes/Vaping | 806 (1.3%) | 1204 (1.3%) |  |
| NA | 173 (0.3%) | 308 (0.3%) |  |
| **Physical activities - past 6 months before test (n, %)** |  |  |  |
| Walk, cycle or light exercise (at least 4 times/week) | 35 920 (58.9%) | 58 848 (64.1%) | < 0.0001 |
| Work out or do gardening (at least 4 times/week) | 15 163 (24.9%) | 18 490 (20.1%) |  |
| Read, watch TV or other sedentary lifestyle | 6742 (11.1%) | 11 696 (12.7%) |  |
| Hard training or competitive sports (several times/week) | 3173 (5.2%) | 2840 (3.1%) |  |
| **Physical form - past 6 months before test (n, %)** |  |  |  |
| Good | 25 003 (41.0%) | 33 536 (36.5%) | < 0.0001 |
| Fair | 21 999 (36.1%) | 37 288 (40.6%) |  |
| Less good | 7010 (11.5%) | 13 109 (14.3%) |  |
| Really good | 5230 (8.6%) | 4602 (5.0%) |  |
| Poor | 1760 (2.9%) | 3343 (3.6%) |  |

Data management and statistical analyses were conducted using R version 4.0.2^22^.

## Results

### Participants

In this study, 430 173 individuals (40.0% test-positive) were invited to complete the questionnaire. A total of 153 412 (35.7%) participants fully completed the questionnaire, 16 125 (3.7%) partially completed the questionnaire, whereas 260 637 (60.7%) individuals were non-responders. The questionnaires were completed approximately 6 (14.7%), 9 (69.7%) and 12 months (15.5%) after the test.

Compared to non-responders, participants who fully completed the baseline questionnaire were more often: females, born in Denmark, older (50-70 years old), more often working within healthcare and living outside of the capital region (table S2).

Among the 171 992 test-positives and 258 181 test-negatives, who were invited to participate, 35.5% and 35.6%, respectively, fully completed the questionnaire, resulting in a total of 152 880 participants after exclusion of 532 test-negatives, who reported having been found seropositive. The participants consisted of 93 494 females (61.2%) and 59 386 males (38.8%) with median ages 50 years (IQRs: 36, 60) and 54 years (IQRs: 41, 64), respectively (table 1). Compared to the test-negatives, test-positives were more often: males, younger, students or having full-time employment, and more physically active, and less often: pensioners or smokers (table 1).

At least one comorbidity were reported by 36.6% of participants (table S2).

### Symptoms around the test date (acute symptoms)

Among test-positives, 84.3% reported at least one “acute” symptom within the period lasting from one week before the test and until four weeks after the test with a median of six symptoms, compared to a median of four among test-negatives with symptoms as test indication. Among all test negatives, irrespective of test indication, 13.5% reported at least one symptom around the test date with a median of two different symptoms. The most common acute symptoms among test-positives were fever (55.0%), fatigue/exhaustion (47.2%) and headache (44.1%) (figure S1). The largest risk differences between test-positives and -negatives tested due to symptoms, were observed for dysgeusia (altered/reduced sense of taste) (RD=34.49%, 95% CI 33.74-35.28%), dysosmia (altered/reduced sense of smell) (RD=33.87%, CI 95% 33.06-34.73%) and fever (RD=23.90%, 95% CI 22.35-25.28%) (figure S1).

### Symptoms 6 to 12 months after test (post-acute symptoms)

Among test-positives, 29.6% reported at least one symptom 6 to 12 months after testing compared to 13.0% of all test-negatives. In both groups, two was the median number of symptoms reported. The three most common symptoms 6 to 12 months after testing-positive were fatigue/exhaustion (11.1%), dysosmia (10.9%) and dysgeusia (8.8%) (figure 1). The most marked risk differences between test-positives and test-negatives 6 to 12 months after test were for: dysosmia (RD=10.92%, 95% CI 10.68-11.21%), dysgeusia (RD=8.68%, 95% CI 8.43-8.93%), and fatigue/exhaustion (RD=8.43%, 95% CI 8.14-8.74%) (figure 1). Additionally, dyspnea, reduced strength in legs/arms, sleeping legs/arms, muscle/joint pain, headache, dizziness, chest pain, reduced appetite, hot flushes/sweat, chills, fever, nausea, diarrhoea, abdominal pain and red runny eyes were all significantly more common among test-positives (figure 1).

**Figure 1:**
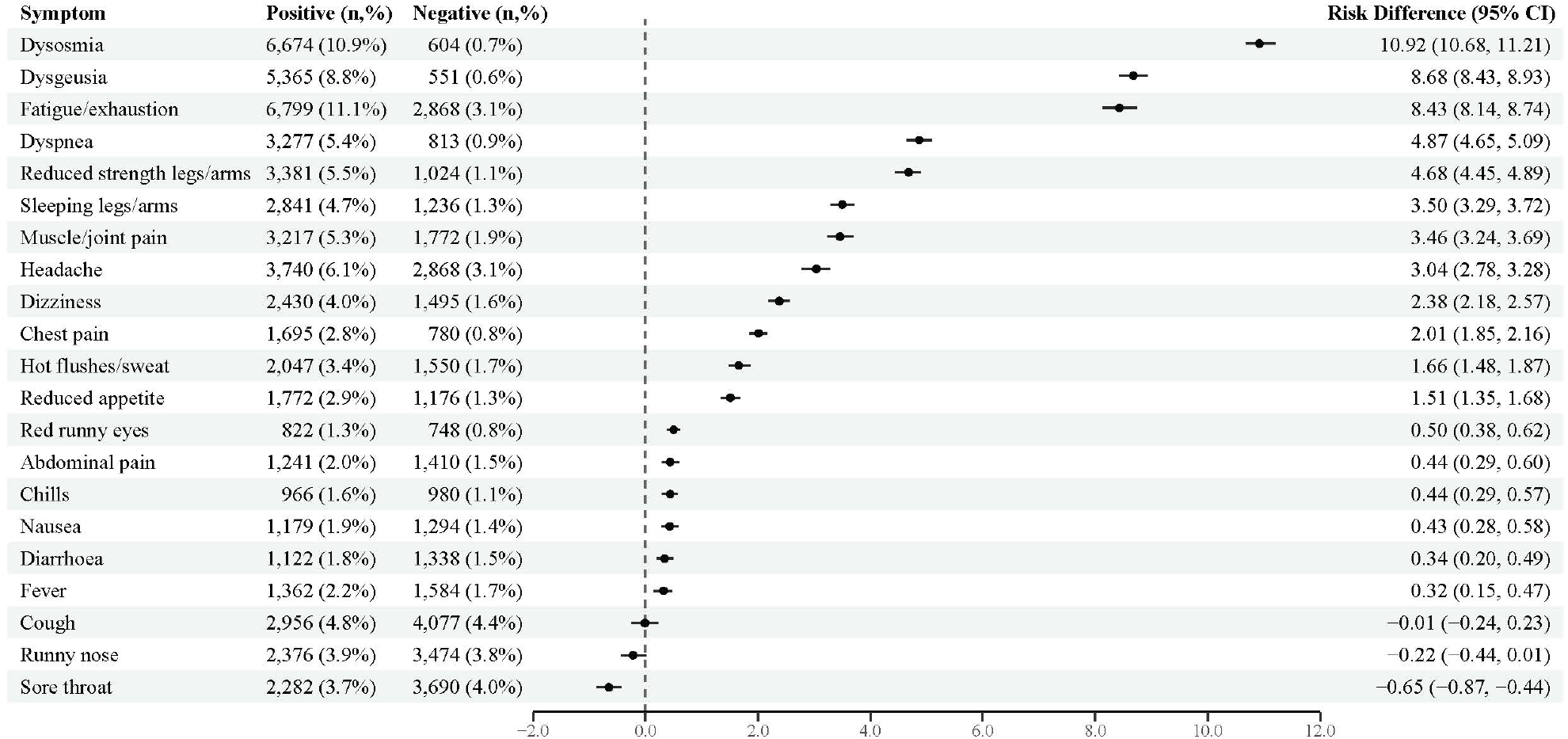
Risk differences of symptoms after 6-12 months, comparing SARS-CoV-2 test-positive and test-negative participants. Note: For post-acute symptoms 6-12 months after the test date, all test-negatives no matter of indication for testing are used as control population. Risk differences with 95% confidence intervals were adjusted for age, sex, comorbidities, obesity and healthcare-occupation.

### New onset of diagnoses and general health problems 6-12 months post test

At least one diagnosis of depression, anxiety, chronic fatigue symptom (CFS), fibromyalgia, or post-traumatic stress disorder (PTSD) with new onset within the first 6, 9 or 12 months after the test was reported by 7.2% of test-positives, compared to 3.3% of test-negatives. The most frequently reported diagnoses were chronic fatigue syndrome (4.0%), depression (3.5%) and anxiety (3.4%) (figure 2). All three diagnoses were more common among test-positives compared to test-negatives with statistically significant risk differences of 2.53% (2.35-2.71%), 1.00% (95% CI 0.81-1.19%) and 1.15% (95% CI 0.95-1.34%), respectively (figure 2). PTSD was also marginally more common among test-positives with a statistically significant risk difference of 0.16% (95% CI 0.03-0.28%).

**Figure 2:**
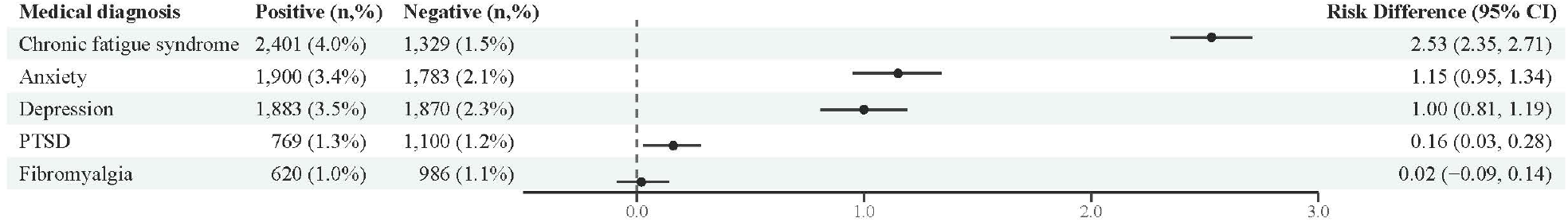
Risk differences of self-reported new diagnoses received between the test date and until 6-12 months after, comparing SARS-CoV-2 test-positive and test-negative participants. Note: For diagnoses with onset up 6-12 months after the test date, all test-negatives no matter of indication for testing are used as control population. Risk differences with 95% confidence intervals were adjusted for age, sex, comorbidities, obesity and healthcare-occupation.

Among test-positives, 53.1% reported at least one of the following problems with new onset within the first 6, 9 or 12 months after the test date: difficulties concentrating; memory problems; mental exhaustion; physical exhaustion or sleep problems, whereas the proportion among test-negatives was 11.5%. The most common problems among test positives were physical exhaustion (RD=40.45%, CI 95% 39.99-40.97%), mental exhaustion (RD=32.58%, 32.11-33.09%), difficulties concentrating (RD=28.34%, CI 95% 27.91-28.78%) and memory issues (RD=27.25%, CI 95% 26.80-27.71%) (figure 3). All the aforementioned health problems were more common among test-positives than test-negatives with large risk differences (figure 3).

**Figure 3:**
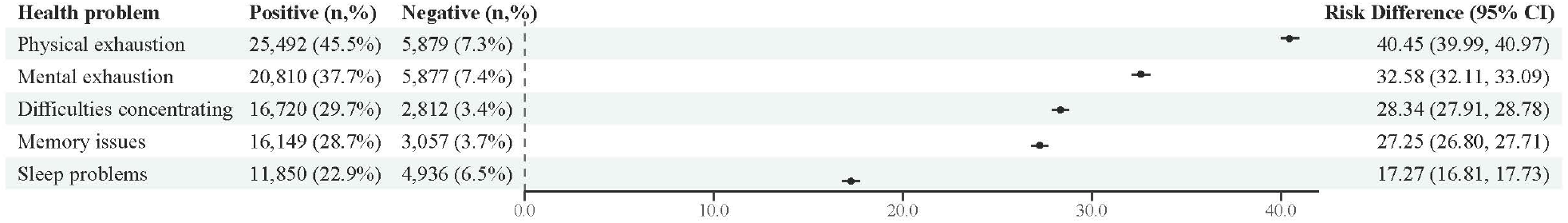
Risk differences of self-reported health problems with new onset between the test date and until 6-12 months after, comparing SARS-CoV-2 test-positive and test-negative participants. Note: For health problems with onset up 6-12 months after the test date, all test-negatives no matter of indication for testing are used as control population. Risk differences with 95% confidence intervals were adjusted for age, sex, comorbidities, obesity and healthcare-occupation.

### Duration of individual symptoms

When looking at estimated RDs for questionnaires completed at 6, 9 or 12 months separately, RDs tended to decrease over time. Among the ten symptoms with largest overall RDs, the estimates decreased over time for all except dysosmia and dysgeusia for which estimates were largest at 9 months (table S3).

### Post-acute symptoms among hospitalized patients

The occurrence of post-acute symptoms among test-positives hospitalized due to covid-19 (4.0%) and non-hospitalized test-positive individuals (96.0%) was compared (figure S2). Considerable risk differences were observed for fatigue/exhaustion (RD=8.64%, 95% CI 6.70-10.74%), reduced strength in arms/legs (RD=7.13%, 95% CI 5.55-8.66%) and dyspnea (RD=6.71%, 95% CI 5.17-8.39). The risk for all symptoms, except for dysgeusia, dysosmia and runny nose were higher among hospitalized than non-hospitalized individuals.

### Post-acute symptoms stratified by age and sex

Risk differences for symptoms 6-12 months after the test were stratified by age group and sex in order to assess the existence of subgroups at greater risk of post-acute symptoms (figure 4, table S4). Based on descriptive results, the majority of post-acute symptoms tended to more often be reported by females and especially by 30-59 year old participants. Stratified RDs for experiencing at least one of the symptoms: fatigue/exhaustion, dysgeusia, dysosmia, 6-12 months after test, were higher for females (RD=18.0 %, 95% CI 17.5 - 18.5%) compared to males (RD=13.1%, 95% CI 12.6 - 13.5%). Additionally, RDs for experiencing at least one of these symptoms were higher for 30-59 year olds (RD=18.2%, 95% CI 17.7 - 18.7%) compared to for all other age groups (15-29 and 60+) (RD=13.5%, 95% CI 13.0 - 13.9%).

**Figure 4:**
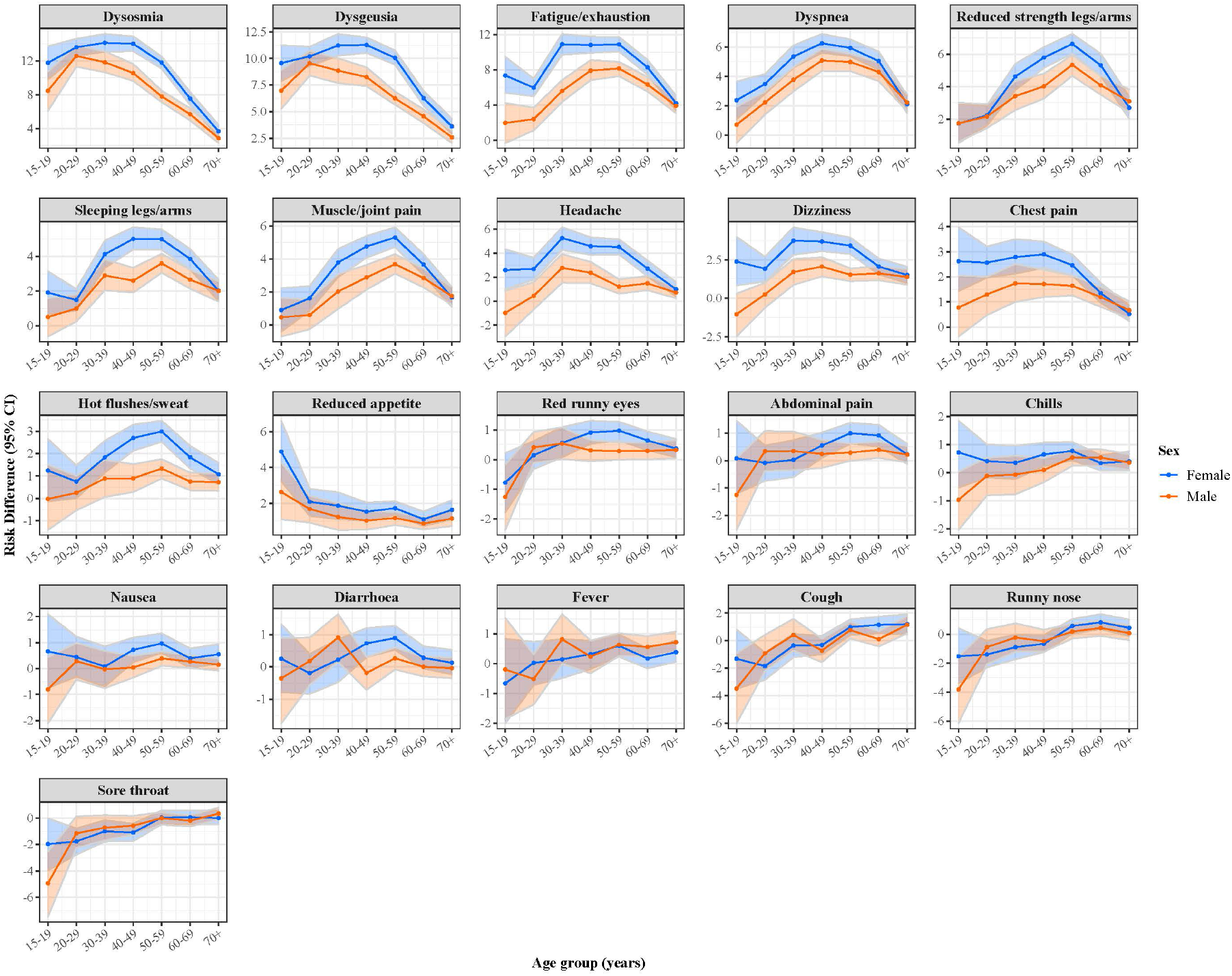
Risk differences of symptoms after 6-12 months, comparing SARS-CoV-2 test-positive and test-negative participants, stratified by sex and age group.

Similar trends and more pronounced differences were observed for new onset of memory-, concentration- or sleep problems, as well as mental or physical exhaustion (figure S3, table S5). Risk differences for new onset of diagnoses of anxiety were highest among 20-29 years old females (figure S4, table S6). Depression were more often reported by 30-39 year olds regardless of sex.

### Sick leave

Full or part-time sick leave was more common after a SARS-CoV-2 infection. Among the test-positives 12.0% reported taking any sick leave 4 weeks after test and until filling in the questionnaire 6-12 months later, compared to 7.7% of test-negatives (RD=4.32%, 95% CI 4.00-4.64%). Full-time sick leave was reported by 9.4% of test-positives and 6.5% of test-negatives (RD=3.20, 95% CI 2.88-3.47%), whereas part-time sick-leave was reported by 4.2% of test-positives compared to 1.7% of test-negatives (RD=2.43%, 95% CI 2.25-2.62%).

## Discussion

Individuals testing positive for SARS-CoV-2 in Denmark during the study period, more often reported physical symptoms, new-onset diagnoses and other health problems 6-12 months after test, compared to test-negative individuals. In particular, there was a marked over-representation of self-reported physical and mental exhaustion among the test-positives, as well as difficulties concentrating, memory issues and sleep problems. New diagnoses of CFS, depression and anxiety were also more common after testing positive. The highest risk differences for physical symptoms were observed for dysosmia, dysgeusia, fatigue/exhaustion and dyspnea. This is consistent with other findings among mainly non-hospitalized patients^11–13,23,24^.

### Other studies

Differences in included symptoms, varying follow-up times, and lack of control groups make direct comparisons of studies difficult. However, in a systematic review, the median prevalence of anosmia and dysgeusia were 11% (IQR, 5.7%-14.3%, 19 studies) and 9% (IQR, 3.0-11.2%, 13 studies)^4^, respectively, which is similar to the present study. In a meta-analysis, the pooled proportion of individuals experiencing fatigue at least 12 weeks after diagnosis, was 32% (95% CI 27, 37)^25^. In the present study, 11.1% of test-positives reported fatigue/exhaustion within the past 14 days, when asked 6-12 months after test, whereas physical or mental exhaustion in general during the time since the test was reported by 45.5% and 37.7%, respectively. Generally, the reported symptom prevalences in our study are in the lower range compared to other studies. However, our study has more follow-up time and is more representative of a general population where the majority of individuals have experienced milder disease. Thus, we believe that our study has greater external validity than many previous studies conducted in hospitalized- or otherwise selected populations.

It is well-established that psychiatric sequelae in the form of anxiety, depression, cognitive problems and sleep disturbances may occur following COVID-19, but reported prevalences vary considerably^26^. Our results suggest that these problems are also prevalent among non-hospitalized individuals^27,28^. The overrepresentation of CFS among test-positives must be interpreted with care due to variability in how this diagnosis is made and the risk of confusing CFS with other conditions when filling in the questionnaire. However, increased incidence of CFS after COVID-19 have also been reported elsewhere^29^.

The number of studies among non-hospitalized individuals with follow-up beyond 6 months are still limited. In one study including 794 test-positive individuals, no specific time gradient were observed in self-rated health 3-8 months post-infection^14^. Others have concluded that recovery beyond 6 months of illness was rare^15^. In the present study, a slightly decreasing trend in reporting frequency at 6, 9 or 12 months was observed for most symptoms.

Increased frequency of post-acute symptoms in females compared to males and slower recovery in females have also been reported in other studies^11–13,15,16^, whereas the evidence regarding the influence of age is somewhat contradicting. In one study, an inverted-U formed association between age and worsening of health after infection was observed, similar to the present study, where the majority of symptoms were most frequently reported by the middle-aged (30-59 years)^14^, but reports of increased risk in older individuals^12,28^, young adults^17^, or no effect^11^ also exist.

The reported differences in sick-leave among test-positives and test-negatives indicate that post-acute symptoms are of such severity that they result in absence from work.

### Strengths and limitations

The main strengths of the present study is its considerable size and the use of a large time-matched control population, making it possible to compare post-acute symptoms among COVID-19 cases and the background population represented by the control group. In addition, we were able to adjust for important confounders, including comorbidity. This allowed us to calculate adjusted risk difference measures for each acute and post-acute symptom, thus ‘deducting’ the general morbidity in the population, including any general health effects that may have been caused by the lock-down or other societal restrictions put in place as part of the epidemic control.

The main limitations of the study are the self-reporting of symptoms and the participation rate. With little over 1/3 of the invitees choosing to participate, we cannot rule out participation bias. The motivation for participation could be higher among those experiencing post-acute symptoms, but on the other hand, those with very severe symptoms might not have had the energy to participate. Still, response rates among test-positives and –negatives were similar. However, because of the size of the study and the marked risk differences between the case- and control groups, we believe that our results are valid.

The current study does not include patient register data, other than data used for calculation of the Charlson comorbidity index scores and defining hospitalizations, meaning that we were not able to address rare, but severe sequelae not caught by the questionnaire. However, results of a study based on the Danish prescription, patient, and health insurance registries, indicated that non-hospitalized patients were at increased risk of being diagnosed with dyspnea and venous thromboembolism, but not other diagnoses^30^.

Our study population were restricted to persons having an e-Boks, which is mandatory, unless exempted, e.g. based on reduced cognitive/physical function, linguistic difficulties, or lack of permanent address. This could potentially introduce bias, but the same groups would most likely also be underrepresented if self-reported data had been collected by other means.

Participants received the questionnaire 6-12 months after the test date of reference, and thus recall bias when reporting symptoms around this date cannot be excluded. Participants were asked to answer to the best of their ability, since we believe that symptoms, which have had an impact on their daily life, would most likely be remembered.

In other to minimize potential influence of recall bias on the reporting of post-acute symptoms, only symptoms experienced within the 14 days up to filling in the questionnaire, were included. For diagnoses made by a doctor or more general problems, we deemed it reasonable to ask for the entire period elapsed since the test date.

## Conclusion

The burden of self-reported symptoms, diagnoses and health issues after SARS-CoV-2 infection appears to be significant in the Danish population and we believe the results are generalizable to other comparable populations. This should be taken into account, when evaluating the full impact of the pandemic and when evaluating the benefits of public health interventions aimed at preventing the spread of the virus.

Further research is needed to better understand, who is at increased risk of developing post-acute disease. Ongoing longitudinal studies are needed to provide more details, particularly on sustained mental health, fatigue and cognitive problems, which this study found to be significantly more often reported among former COVID-19 patients than controls.

## Supporting information

STROBE check list

## Data Availability

Health data are considered person-sensitive, and cannot be shared due to data protection regulations.

## Author contributions

The study was designed and initiated by: PB, SE, AH, NN, AK, AS

The questionnaire was designed by: AS, PB, NN, AK, SE, IM

Collection of data including programming related to this: AS, JH, PB, IM

Data analysis was done by: LS

The first draft was written by: AS, LS, AH

All authors have critically revised the manuscript.

All authors have approved the final version of the manuscript and agreed to be accountable for all aspects of the work.

## Acknowledgements

The authors would like to thank all EFTER-COVID participants for filling in the questionnaire.

We also thank, the members of the EFTER-COVID stakeholder group, consisting of representatives from universities, long-term sequelae clinics and other relevant hospital departments or health institutions, for their comments to the questionnaires and input during meetings.

## References

1 Marshall M. The lasting misery of coronavirus long-haulers. Nature 2020; 585: 339–41.

2 World Health Organization. A clinical case definition of post COVID-19 condition by a Delphi consensus. 2021 https://www.who.int/publications/i/item/WHO-2019-nCoV-Post_COVID-19_condition-Clinical_case_definition-2021.1.

3 Soriano JB, Murthy S, Marshall JC, Relan P, Diaz J V. A clinical case definition of post-COVID-19 condition by a Delphi consensus. Lancet Infect Dis 2021; 3099: 19–24.

4 Nasserie T, Hittle M, Goodman SN. Assessment of the Frequency and Variety of Persistent Symptoms among Patients with COVID-19: A Systematic Review. JAMA Netw Open 2021; 4: 1– 19.

5 Groff D, Sun A, Ssentongo AE, et al. Short-term and Long-term Rates of Postacute Sequelae of SARS CoV-2 Infection: A Systematic Review. JAMA Netw open 2021; 4: e2128568.

6 Taquet M, Geddes JR, Husain M, Luciano S, Harrison PJ. 6-month neurological and psychiatric outcomes in 236 379 survivors of COVID-19 : a retrospective cohort study using electronic health records. Lancet Psychiatry 2021. https://doi.org/10.1016/S2215-0366(21)00084-5.

7 Christian HH, Daniela M, Sophie MG, Kåre M, Steen E. Assessment of protection against reinfection with SARS-CoV-2 among 4 million PCR-tested individuals in Denmark in 2020: a population-level observational study Christian. Lancet 2020; 19–21.

8 Statens Serum Institut. COVID-19: Branche fordelte opgørelser over covid-19-testede og positive. https://covid19.ssi.dk/overvagningsdata/branchefordelte-opgoerelser (accessed Jan 25, 2022).

9 Lynge E, Sandegaard JL, Rebolj M. The Danish national patient register. Scand J Public Health 2011; 39: 30–3.

10 Robins JM. A New Approach To Causal Inference in Mortality Studies With a Sustained Exposure Period -Application To Control of the Healthy Worker Survivor Effect’. Math Model 1986; 7: 1393– 512.

11 Bliddal S, Banasik K, Pedersen OB, et al. Acute and persistent symptoms in non-hospitalized PCR-confirmed COVID-19 patients. Sci Rep 2021; 11: 1–11.

12 Augustin M, Schommers P, Stecher M, et al. Post-COVID syndrome in non-hospitalised patients with COVID-19: a longitudinal prospective cohort study. Lancet Reg Heal - Eur 2021; 6: 1–8.

13 Stavem K, Ghanima W, Olsen MK, Gilboe HM, Einvik G. Persistent symptoms 1.5-6 months after COVID-19 in non-hospitalised subjects: A population-based cohort study. Thorax 2021; 76: 405–7.

14 Søraas A, Kalleberg KT, Dahl JA, et al. Persisting symptoms three to eight months after non-hospitalized COVID-19, a prospective cohort study. PLoS One 2021; 16: 1–13.

15 Wynberg E, van Willigen HD, Dijkstra M, et al. Evolution of COVID-19 symptoms during the first 12 months after illness onset. Clin Infect Dis 2021; Sep 2, onl. DOI:10.1093/cid/ciab759.

16 Kashif A, Chaudhry M, Fayyaz T, et al. Follow-up of COVID-19 recovered patients with mild disease. Sci Rep 2021; 11: 1–5.

17 Taquet M, Dercon Q, Luciano S, Geddes JR, Husain M, Harrison PJ. Incidence, co-occurrence, and evolution of long-COVID features: A 6-month retrospective cohort study of 273,618 survivors of COVID-19. PLoS Med 2021; 18: 1–22.

18 Grembi J, McQuade ER. riskCommunicator: G-Computation to Estimate Interpretable Epidemiological Effects. R package version 0.1.0. 2020. https://cran.r-project.org/package=riskCommunicator.

19 Boyes R. Forester: An R package for creating publication-ready forest plots. 2021. https://github.com/rdboyes/forester.

20 Quan H, Sundararajan V, Halfon P, et al. Coding algorithms for defining comorbidities in ICD-9-CM and ICD-10 administrative data. Med Care 2005; 43: 1130–9.

21 Cole TJ, Bellizzi MC, Flegal KM, Dietz WH. Establishing a standard definition for child overweight and obesity Worldwide: international Survey. BMJ 2000; 320: 1–6.

22 R Core team. R: A language and environment for statistical computing. R Foundation for Statistical Computing, Vienna, Austria. 2021. http://www.r-project.org/.

23 [preprint] Rauch B, Kern-Matschilles S, Haschka SJ, et al. COVID-19-related symptoms 6 months after the infection - Update on a prospective cohort study in Germany. medRxiv Prepr Serv Heal Sci 2021; : 1–11.

24 Glück V, Grobecker S, Tydykov L, et al. SARS-CoV-2-directed antibodies persist for more than six months in a cohort with mild to moderate COVID-19. Infection 2021; 49: 739–46.

25 Ceban F, Ling S, Lui LMW, et al. Fatigue and Cognitive Impairment in Post-COVID-19 Syndrome: A Systematic Review and Meta-Analysis. Brain Behav Immun 2021. DOI:10.1016/j.bbi.2021.12.020.

26 Schou TM, Joca S, Wegener G, Bay-Richter C. Psychiatric and neuropsychiatric sequelae of COVID-19 – A systematic review. Brain Behav Immun 2021; 97: 328–48.

27 Al-Aly Z, Xie Y, Bowe B. High-dimensional characterization of post-acute sequelae of COVID-19. Nature 2021; 594: 259–64.

28 Daugherty SE, Guo Y, Heath K, et al. Risk of clinical sequelae after the acute phase of SARS-CoV-2 infection: Retrospective cohort study. BMJ 2021; 373. DOI:10.1136/bmj.n1098.

29 [preprint] Roessler M, Tesch F, Batram M, Jacob J, Loser F, Weidinger O. Post COVID - 19 in children, adolescents, and adults : results of a matched cohort study including more than 150, 000 individuals with COVID - 19. medRxiv Prepr 2021; : 1–19.

30 Lund LC, Hallas J, Nielsen H, et al. Post-acute effects of SARS-CoV-2 infection in individuals not requiring hospital admission: a Danish population-based cohort study. Lancet Infect Dis 2021; 3099: 1–10.

