## Supplementary materials for "Post-acute symptoms, new onset diagnoses and health problems 6 to 12 months after SARS-CoV-2 infection: a nationwide questionnaire study in the adult Danish population"

**Text S1: English translation of the questionnaire**

*Background information:* This is a print of an electronic questionnaire containing activations. This means that when filling out the questionnaire electronically, it depends on test status and/or answers to previous questions, which questions a given participant will be asked next. In this print, activations will be indicated by explanatory text in square brackets. When send out to participants, their specific test date of reference, were inserted in all places indicated by [*test date*]. In one some questions, the listed time intervals differed depending on whether participants received the questionnaire 6, 9 or 12 month after the test. In this example, the intervals for at 6 months are shown.

*[Start page in the questionnaire]*

**Welcome to the EFTER-COVID baseline questionnaire**

In the EFTER-COVID project, we are examining the health of the population during and after the coronavirus pandemic. **Your answers are important, regardless of whether you have tested positive or negative for coronavirus (COVID-19)**

Everyone invited to participate in the study has been randomly selected to receive a questionnaire with a particular focus on a certain area of their health. The questionnaire you have received focuses on physical symptoms.

**Many thanks for your help.**

[*Test-positive participants only*]:

**These first questions are about how you have been since you tested positive at [*testdate*]**

Which of the following conditions best describes how you were when you were at your <u>worst?</u>

Think about the period from a week before you tested positive until 4 weeks after.

(1) ☐ I had no symptoms

(2) ☐ I was ill at home

(3) ☐ I was hospitalised due to COVID-19

(4) ☐ I was hospitalised due to COVID-19 and was intubated with a ventilator

(5) ☐ I was hospitalised for reasons other than COVID-19

[*Test-positive participants only*]:

**Did you take any of the following types of medication for your COVID-19 symptoms?**

Think of the period from one week before you were tested at [*test date*] and until 4 weeks after.

(1) ☐ Painkillers/antipyretics

(2) ☐ Asthma medication

(3) ☐ Other - please specify _____

(4) ☐ I didn't take any medication

[*Test-negative participants only*]:

**Why were you tested on the [*test date*]?**

(1) ☐ I had symptoms

(2) ☐ I was a close contact of someone infected

(3) ☐ I suspected that I had been exposed to the infection

(4) ☐ Other

(5) ☐ I can't remember

**Physical symptoms in the period around the test.**

**Have you at any point had any of the following symptoms?** Please consider the period from the week leading up to your test on [*test date*] and until now.

Kindly mark all the symptoms you have had without considering the symptom cause. You can add further comments at the end of the questionnaire.

| | Yes | No | Don't know |
|---|---|---|---|
| Fever | (1) ☐ | (0) ☐ | (99) ☐ |
| Chills | (1) ☐ | (0) ☐ | (99) ☐ |
| Hot flushes / sweating | (1) ☐ | (0) ☐ | (99) ☐ |
| Headache | (1) ☐ | (0) ☐ | (99) ☐ |
| Muscle or joint pain | (1) ☐ | (0) ☐ | (99) ☐ |
| Fatigue or exhaustion | (1) ☐ | (0) ☐ | (99) ☐ |
| Dizziness | (1) ☐ | (0) ☐ | (99) ☐ |
| Red or watery eyes | (1) ☐ | (0) ☐ | (99) ☐ |
| Runny or stuffy nose | (1) ☐ | (0) ☐ | (99) ☐ |

**Continued from previous page: In the week leading up to the test on [*test date*], and until now, did you have any of the following symptoms?**

Kindly mark all the symptoms you have had without considering the symptom cause. You can add further comments at the end of the questionnaire.

| | Yes | No | Don't know |
|---|---|---|---|
| Reduced or altered sense of smell | (1) ☐ | (0) ☐ | (99) ☐ |
| Reduced or altered sense of taste | (1) ☐ | (0) ☐ | (99) ☐ |
| Cough | (1) ☐ | (0) ☐ | (99) ☐ |
| Sore throat or pain when swallowing | (1) ☐ | (0) ☐ | (99) ☐ |
| Shortness of breath | (1) ☐ | (0) ☐ | (99) ☐ |
| Chest pain | (1) ☐ | (0) ☐ | (99) ☐ |
| Nausea or vomiting | (1) ☐ | (0) ☐ | (99) ☐ |
| Diarrhoea | (1) ☐ | (0) ☐ | (99) ☐ |
| Abdominal pain | (1) ☐ | (0) ☐ | (99) ☐ |
| Reduced or loss of appetite | (1) ☐ | (0) ☐ | (99) ☐ |
| Reduced arm and leg strength | (1) ☐ | (0) ☐ | (99) ☐ |
| Sleeping or tingling sensation, or other abnormal sensations in the legs and arms | (1) ☐ | (0) ☐ | (99) ☐ |
| Other symptoms | (1) ☐ | (0) ☐ | (99) ☐ |

[*In this question, only lines related to symptoms, which the participants have replied "yes" to in the previous question are displayed*]

**During which periods did you have symptoms?**

**"Around the test date" covers the period from a week before the test on [*test date*] and up to four weeks after.**

Please mark all of the periods during which you have had the symptom. If you have trouble remembering, then please mark the periods to the best of your ability.

| | Around the test date | 1-2 months following the test | 2-6 months following the test | The past 14 days |
|---|---|---|---|---|
| Fever | (1) ☐ | (12) ☐ | (26) ☐ | (14) ☐ |
| Chills | (1) ☐ | (12) ☐ | (26) ☐ | (14) ☐ |
| Hot flushes / sweating | (1) ☐ | (12) ☐ | (26) ☐ | (14) ☐ |
| Headache | (1) ☐ | (12) ☐ | (26) ☐ | (14) ☐ |
| Muscle or joint pain | (1) ☐ | (12) ☐ | (26) ☐ | (14) ☐ |
| Fatigue or exhaustion | (1) ☐ | (12) ☐ | (26) ☐ | (14) ☐ |
| Dizziness | (1) ☐ | (12) ☐ | (26) ☐ | (14) ☐ |
| Red or watery eyes | (1) ☐ | (12) ☐ | (26) ☐ | (14) ☐ |
| Runny or stuffy nose | (1) ☐ | (12) ☐ | (26) ☐ | (14) ☐ |
| Reduced or altered sense of smell | (1) ☐ | (12) ☐ | (26) ☐ | (14) ☐ |

| | | | | |
|---|---|---|---|---|
| Reduced or altered sense of taste | (1) ☐ | (12) ☐ | (26) ☐ | (14) ☐ |
| Sore throat or pain when swallowing | (1) ☐ | (12) ☐ | (26) ☐ | (14) ☐ |
| Cough | (1) ☐ | (12) ☐ | (26) ☐ | (14) ☐ |
| Shortness of breath | (1) ☐ | (12) ☐ | (26) ☐ | (14) ☐ |
| Chest pain | (1) ☐ | (12) ☐ | (26) ☐ | (14) ☐ |
| Nausea or vomiting | (1) ☐ | (12) ☐ | (26) ☐ | (14) ☐ |
| Diarrhoea | (1) ☐ | (12) ☐ | (26) ☐ | (14) ☐ |
| Abdominal pain | (1) ☐ | (12) ☐ | (26) ☐ | (14) ☐ |
| Reduced or loss of appetite | (1) ☐ | (12) ☐ | (26) ☐ | (14) ☐ |
| Reduced arm and leg strength | (1) ☐ | (12) ☐ | (26) ☐ | (14) ☐ |
| Sleeping or tingling sensation, or other abnormal sensations in the legs and arms | (1) ☐ | (12) ☐ | (26) ☐ | (14) ☐ |

[*Only participants, who replied "yes" to having had fever*]:

**Did you take your temperature when you had a fever in the period around your test on the [*test date*]?**

(1) ☐ Yes (please write the highest temperature taken in the box below) _____

(0) ☐ No

(99) ☐ Don't know/Can't remember

[*Only participants, who replied "yes" to having had fever*]:

**Were there known/probable causes other than COVID-19 for you having a fever in the period around the test on [*test date*]?**

(1) ☐ Yes (please explain in the box below) _____

(0) ☐ No

(99) ☐ Don't know

[*In this question, only lines related to symptoms, which the participants have replied "yes" to having experienced, will be displayed*]

**Before the test on [*test date*], did you generally (i.e. often or chronically) suffer from any of the following physical symptoms?**

| | Yes | No | Don't know |
|---|---|---|---|
| Fever | (1) ☐ | (0) ☐ | (99) ☐ |
| Chills | (1) ☐ | (0) ☐ | (99) ☐ |
| Hot flushes / sweating | (1) ☐ | (0) ☐ | (99) ☐ |
| Headache | (1) ☐ | (0) ☐ | (99) ☐ |
| Muscle or joint pain | (1) ☐ | (0) ☐ | (99) ☐ |
| Fatigue or exhaustion | (1) ☐ | (0) ☐ | (99) ☐ |
| Dizziness | (1) ☐ | (0) ☐ | (99) ☐ |
| Red or watery eyes | (1) ☐ | (0) ☐ | (99) ☐ |

| | | | |
|---|---|---|---|
| Runny or stuffy nose | (1) ☐ | (0) ☐ | (99) ☐ |
| Reduced or altered sense of smell | (1) ☐ | (0) ☐ | (99) ☐ |
| Reduced or altered sense of taste | (1) ☐ | (0) ☐ | (99) ☐ |
| Sore throat or pain when swallowing | (1) ☐ | (0) ☐ | (99) ☐ |
| Cough | (1) ☐ | (0) ☐ | (99) ☐ |
| Shortness of breath | (1) ☐ | (0) ☐ | (99) ☐ |
| Chest pain | (1) ☐ | (0) ☐ | (99) ☐ |
| Nausea or vomiting | (1) ☐ | (0) ☐ | (99) ☐ |
| Diarrhoea | (1) ☐ | (0) ☐ | (99) ☐ |
| Abdominal pain | (1) ☐ | (0) ☐ | (99) ☐ |
| Reduced or loss of appetite | (1) ☐ | (0) ☐ | (99) ☐ |
| Reduced arm and leg strength | (1) ☐ | (0) ☐ | (99) ☐ |
| Sleeping or tingling sensation, or other abnormal sensations in the legs and arms | (1) ☐ | (0) ☐ | (99) ☐ |

[*Only participants, who replied "yes" to having had headache. Only lines representing time intervals, where participants indicated having headache are displayed*]:

**How severe were your headaches when you were at your worst during the specified period(s)?**

| | No pain | Mild pain | Moderate pain | Strong pain | Unbearable pain |
|---|---|---|---|---|---|
| Generally before the test date | (1) ☐ | (2) ☐ | (3) ☐ | (4) ☐ | (5) ☐ |
| Around the test date | (1) ☐ | (2) ☐ | (3) ☐ | (4) ☐ | (5) ☐ |
| 1-2 months following the test | (1) ☐ | (2) ☐ | (3) ☐ | (4) ☐ | (5) ☐ |
| 2-6 months following the test | (1) ☐ | (2) ☐ | (3) ☐ | (4) ☐ | (5) ☐ |
| The past 14 days | (1) ☐ | (2) ☐ | (3) ☐ | (4) ☐ | (5) ☐ |

[*Only participants, who replied "yes" to having had muscle or joint pain. Only lines representing time intervals, where participants indicated having this are displayed*]:

**How bad was your muscle or joint pain in the specified period(s)?**

| | No pain | Mild pain | Moderate pain | Strong pain | Unbearable pain |
|---|---|---|---|---|---|
| Generally before the test date | (1) ☐ | (2) ☐ | (3) ☐ | (4) ☐ | (5) ☐ |
| Around the test date | (1) ☐ | (2) ☐ | (3) ☐ | (4) ☐ | (5) ☐ |
| 1-2 months following the test | (1) ☐ | (2) ☐ | (3) ☐ | (4) ☐ | (5) ☐ |
| 2-6 months following the test | (1) ☐ | (2) ☐ | (3) ☐ | (4) ☐ | (5) ☐ |
| The past 14 days | (1) ☐ | (2) ☐ | (3) ☐ | (4) ☐ | (5) ☐ |

[*Only participants, who replied "yes" to having had reduced or changed sense of smell. Only lines representing time intervals, where participants indicated having this are displayed*]:

**How was your sense of smell in the specified period(s), compared to normal?**

My sense of smell was/is…

| | Slightly reduced | Very reduced | Completely gone | Changed | Don't know |
|---|---|---|---|---|---|
| Generally before the test date | (1) ☐ | (2) ☐ | (3) ☐ | (4) ☐ | (5) ☐ |
| Around the test date | (1) ☐ | (2) ☐ | (3) ☐ | (4) ☐ | (5) ☐ |
| 1-2 months following the test | (1) ☐ | (2) ☐ | (3) ☐ | (4) ☐ | (5) ☐ |
| 2-6 months following the test | (1) ☐ | (2) ☐ | (3) ☐ | (4) ☐ | (5) ☐ |
| The past 14 days | (1) ☐ | (2) ☐ | (3) ☐ | (4) ☐ | (5) ☐ |

[*Only participants, who replied "yes" to having had reduced or changed sense of taste. Only lines representing time intervals, where participants indicated having this are displayed*]:

**How was your sense of taste in the specified period(s), compared to normal?**

My sense of taste was/is…

| | Slightly reduced | Very reduced | Completely gone | Changed | Don't know |
|---|---|---|---|---|---|
| Generally before the test date | (1) ☐ | (2) ☐ | (3) ☐ | (4) ☐ | (5) ☐ |
| Around the test date | (1) ☐ | (2) ☐ | (3) ☐ | (4) ☐ | (5) ☐ |
| 1-2 months following the test | (1) ☐ | (2) ☐ | (3) ☐ | (4) ☐ | (5) ☐ |

| | | | | | |
|---|---|---|---|---|---|
| 2-6 months following the test | (1) ☐ | (2) ☐ | (3) ☐ | (4) ☐ | (5) ☐ |
| The past 14 days | (1) ☐ | (2) ☐ | (3) ☐ | (4) ☐ | (5) ☐ |

[*Only participants, who replied "yes" to having experienced shortness of breath*. *Only lines representing time intervals, where participants indicated having this are displayed*]:

**Choose the following statement below that best describes your shortness of breath during the relevant period(s).**

**A. I only become short of breath when I exert myself a lot**

**B. I become short of breath when I hurry or walk up a small hill**

**C. I walk slower than others my age due to my shortness of breath, or I have to stop to catch my breath when walking on even terrain**

**D. I stop to catch my breath after about 100 metres or a few minutes of walking**

**E. I have too much shortness of breath to leave my home, or I get short of breath when getting (un)dressed**

| | A | B | C | D | E | Don't know |
|---|---|---|---|---|---|---|
| Generally before the test date | (1) ☐ | (2) ☐ | (3) ☐ | (4) ☐ | (5) ☐ | (99) ☐ |
| Around the test date | (1) ☐ | (2) ☐ | (3) ☐ | (4) ☐ | (5) ☐ | (99) ☐ |
| 1-2 months following the test | (1) ☐ | (2) ☐ | (3) ☐ | (4) ☐ | (5) ☐ | (99) ☐ |
| 2-6 months following the test | (1) ☐ | (2) ☐ | (3) ☐ | (4) ☐ | (5) ☐ | (99) ☐ |
| In the past 14 days | (1) ☐ | (2) ☐ | (3) ☐ | (4) ☐ | (5) ☐ | (99) ☐ |

*[Only participants indicating having had other symptoms]:*

**Which other symptoms have you had and when?**

________________________________________
________________________________________
________________________________________
________________________________________
________________________________________
________________________________________

*[Only participants indicating having had other symptoms]:*

**Did you also used to have these symptoms generally (i.e. chronically or often) before the test on [*test date*]?**

________________________________________
________________________________________
________________________________________
________________________________________
________________________________________
________________________________________

[*test-positives only*]:

**Did your health return to normal within 4 weeks of testing positive for COVID-19?**

(0) No, my health at this time was not yet completely back to as it was before I became ill

(1) Yes, my health returned to normal a number of days after symptoms started (write the number of days in the field below) _____

(99) Don't know

**Did you take sick leave around the time of the test on [*test date*], or any time since then?**

Please mark all of the relevant answers, regardless of the cause of illness

(1) Yes, I was on sick leave in the week up to the test or up to 4 weeks after

(2) Yes, I was on sick leave more than 4 weeks after the test

(0) No

(3) Not relevant

[*Only shown for participants, who indicated taking sick leave around the test date*]:

**How long were you on sick leave in the period around the test?**

Think about the period from one week before the test on [test date] and up to 4 weeks after.

Please write the number of sick days in the box below. If you can't remember the exact number, write an approximate amount of days. Have you not been on sick leave, please write '0' in the boxes below.

Full-time sick leave:

_____

Part-time sick leave:

_____

[*Only shown for participants, who indicated taking sick leave later than 4 weeks after the test date*]:

**If you were on sick leave later than 4 weeks after the test date, were you on full-time or part-time sick leave?**

If you have been on both full-time and part-time sick leave, please mark both answers below

(1) ☐ Full-time sick leave

(2) ☐ Part-time sick leave

[*test-positives only*]:

**Was your sick leave related to you having had COVID-19?**

(1) ☐ Yes

(0) ☐ No

(2) ☐ Partly

(99) ☐ Don't know

[*Only participants, who indicated having been on full-time sick leave*]:

**How long were you on full-time sick leave?**

(2) ☐ Less than 2 weeks

(4) ☐ 2-4 weeks

(12) ☐ 1-2 months

(24) ☐ 2-4 months

(46) ☐ 4-6 months

(10,000) ☐ I have been on full-time sick leave since the test date

[*Only participants, who indicated having been on part-time sick leave*]:

**How long were you on part-time sick leave?**

(2) □ Less than 2 weeks

(4) □ 2-4 weeks

(12) □ 1-2 months

(24) □ 2-4 months

(46) □ 4-6 months

(10,000) □ I have been on part-time sick leave since the test date

[*Only participants, who replied indicated having been on sick leave*]:

**Has any of your sick leave been within the past 14 days?**

(1) □ Yes

(0) □ No

(3) □ Not relevant

The following pages contain some background questions about your health, education, work and lifestyle.

**Please provide your height in cm**

If you don't know your height, or do not wish to answer, please click "Next" below.

_____

**Please provide your approximate weight in kg.**

If you don’t know your weight, or do not wish to answer, please click “Next” below

_____

**What is your highest level of education?**

(1) □ Primary/Elementary school (9th–10th grade)

(2) □ General secondary education or vocational secondary education

(3) □ Vocational training

(4) □ Shorter term higher education (1–2 years), e.g. vocational academy

(5) □ Medium term higher education (2–4 years), e.g. nursing, primary school teaching, BSc

(6) □ Longer term higher education (more than 5 years), e.g. Masters or PhD

(99) □ Don’t know/None of the above/Do not wish to answer

**What is your main occupational status?**

(1) ☐ Employed full-time

(2) ☐ Employed part-time

(3) ☐ Self-employed

(4) ☐ Student

(5) ☐ Stay-at-home parent or on parental leave

(6) ☐ Jobseeking/Unemployed

(7) ☐ Benefits recipient

(8) ☐ Long-term sick leave

(9) ☐ Pensioner or early retiree

(10) ☐ Other

**Do you smoke or have you previously smoked?**

(1) ☐ I've never smoked

(2) ☐ It has been more than 5 years since I smoked

(3) ☐ I have smoked within the past 5 years, but no longer smokes

(4) ☐ I smoke occasionally, e.g. social events

(5) ☐ I smoke daily (fewer than 10 cigarettes/cigars/pipes per day)

(6) ☐ I smoke daily (10 or more cigarettes/cigars/pipes per day)

(7) ☐ I smoke e-cigarettes/I vape

**Please indicate how many alcoholic drinks you have on average per week.**

One drink is for example 1 beer (330ml), 1 glass of wine (120ml), 1 glass of fortified wine (80ml), or 1 shot of spirits (40ml).

If you don't know, or don't wish to answer, please click "Next" below.

_____

**If you look at the six months before the test on [*test date*], what would you say best describes your level of physical activity in your spare time?**

(1) ☐ Train hard and play competitive sports regularly, several times a week

(2) ☐ Work out or do heavy gardening or similar at least 4 times a week

(3) ☐ Walk, cycle or other light exercise at least 4 times a week (including leisure walks, light gardening, and cycling/walking to work)

(4) ☐ Read, watch TV or other sedentary lifestyle

**If you look at the six months before the test on [*test date*], how would you describe your physical fitness?**

(1) ☐ Really good

(2) ☐ Good

(3) ☐ Fair

(4) ☐ Less good

(5) ☐ Poor

**In the period six months before the test on [*test date*] and up to now, have you had any of the following problems?**

Please mark all the relevant answers

| | Yes, before the test date | Yes, after the test date | No | Don't know |
|---|---|---|---|---|
| Difficulties concentrating | (1) ☐ | (2) ☐ | (0) ☐ | (99) ☐ |
| Issues with memory | (1) ☐ | (2) ☐ | (0) ☐ | (99) ☐ |
| Mental exhaustion | (1) ☐ | (2) ☐ | (0) ☐ | (99) ☐ |
| Physical exhaustion | (1) ☐ | (2) ☐ | (0) ☐ | (99) ☐ |
| Sleep problems | (1) ☐ | (2) ☐ | (0) ☐ | (99) ☐ |

**Have you ever been diagnosed by a doctor with any of the following conditions?**

| | Yes, before the test date | Yes, after the test date | No |
|---|---|---|---|
| Depression | (1) ☐ | (2) ☐ | (0) ☐ |
| Anxiety | (1) ☐ | (2) ☐ | (0) ☐ |
| PTSD (Post traumatic stress disorder) | (1) ☐ | (2) ☐ | (0) ☐ |
| Chronic fatigue syndrome | (1) ☐ | (2) ☐ | (0) ☐ |
| Fibromyalgia | (1) ☐ | (2) ☐ | (0) ☐ |

**Before the test on [*test date*], were you diagnosed with any chronic disease(s)?**

Please mark all relevant.

(1) ☐ No known chronic diseases

(2) ☐ Diabetes

(3) ☐ Asthma

(4) ☐ High blood pressure

(5) ☐ COPD or other chronic lung disease

(6) ☐ Chronic or frequent headaches, including migraines

(7) ☐ Other chronic diseases

(8) ☐ Do not wish to respond

[*Test-negatives only*]:

**Do you think that you have ever had covid-19?**

(1) ☐ Yes

(0) ☐ No

(99) ☐ Don't know

[*Test-negatives, replying yes to the previous question*]:

**Why do you think you probably had covid-19?**

Please mark all relevant.

(1) ☐ Antibodies were detected in my blood sample

(2) ☐ I had symptoms and I think it was covid-19

(3) ☐ I had symptoms, and talked to a doctor who thought it was covid-19

(4) ☐ For other reasons

**Many thanks for taking the time to complete our questionnaire.**

**May we contact you again?**

(1) ☐ Yes

(0) ☐ No

**If you have any further comments, feel free to write them in the box below.**

Please, remember to click "Finish"

____________________________________________

____________________________________________

____________________________________________

____________________________________________

____________________________________________

____________________________________________

**Table S1: Characteristics of the invited persons, stratified by participation status**

| | Positive | Negative | p-value |
|---|---|---|---|
| **Age group (n,%)** | | | |
| 15-19 | 2350 (3.9%) | 3181 (3.5%) | < 0.0001 |
| 20-29 | 9021 (14.8%) | 8958 (9.7%) | |
| 30-39 | 7979 (13.1%) | 9838 (10.7%) | |
| 40-49 | 11 377 (18.7%) | 16 117 (17.5%) | |
| 50-59 | 14 946 (24.5%) | 23 873 (26.0%) | |
| 60-69 | 9530 (15.6%) | 18 975 (20.7%) | |
| 70+ | 5799 (9.5%) | 10 936 (11.9%) | |
| **Obesity (n,%)** | | | |
| Non-obese | 46 256 (75.8%) | 68 712 (74.8%) | < 0.0001 |
| Obese | 9973 (16.3%) | 15 073 (16.4%) | |
| No information | 4773 (7.8%) | 8093 (8.8%) | |
| **Charlson comorbidity score (n,%)** | | | |
| 0 | 53 530 (87.8%) | 78 694 (85.7%) | < 0.0001 |
| 1 | 3807 (6.2%) | 6309 (6.9%) | |
| 2 or more | 3665 (6.0%) | 6875 (7.4%) | |
| **Region (n, %)** | | | |
| Hovedstaden | 26 242 (43.0%) | 29 768 (32.4%) | < 0.0001 |
| Midtjylland | 11 233 (18.4%) | 19 737 (21.5%) | |
| Syddanmark | 9557 (15.7%) | 19 134 (20.8%) | |
| Sjælland | 9233 (15.1%) | 12 678 (13.8%) | |
| Nordjylland | 4706 (7.7%) | 10 504 (11.4%) | |
| No information | 31 (0.1%) | 57 (0.1%) | |
| **Origin (n, %)** | | | |
| Danish | 52 622 (86.3%) | 84 045 (91.5%) | < 0.0001 |
| Born abroad | 7122 (11.7%) | 7165 (7.8%) | |
| Immigrant | 1257 (2.1%) | 665 (0.7%) | |
| **Comorbidities (n, %)** | | | |
| Diabetes | 2647 (4.3%) | 4202 (4.6%) | 0.03 |
| Asthma | 4579 (7.5%) | 6167 (6.7%) | < 0.0001 |
| High blood pressure | 8842 (14.5%) | 14 903 (16.2%) | < 0.0001 |
| COPD or other chronic lung disease | 1187 (1.9%) | 2453 (2.7%) | < 0.0001 |
| Chronic headache (including migraine) | 2138 (3.5%) | 3457 (3.8%) | 0.01 |
| Other chronic disease | 7968 (13.1%) | 14 751 (16.1%) | < 0.0001 |
| **Occupation (n, %)** | | | |
| Non-healthcare worker | 54 400 (89.2%) | 79 654 (86.7%) | < 0.0001 |
| Healthcare worker | 6602 (10.8%) | 12 224 (13.3%) | |

**Table S2: Characteristics of participants, stratified by participation**

| | Non-participants | Full-participants | Partial-participants | p-value |
|---|---|---|---|---|
| **Test result (n,%)** | | | | |
| Negative | 154 085 (59.1%) | 91 878 (60.1%) | 11 687 (72.5%) | < 0.0001 |
| Positive | 106 552 (40.9%) | 61 002 (39.9%) | 4438 (27.5%) | |
| **Sex (n, %)** | | | | |
| Female | 133 489 (51.2%) | 93 494 (61.2%) | 10 246 (63.5%) | < 0.0001 |
| Male | 127 148 (48.8%) | 59 386 (38.8%) | 5879 (36.5%) | |
| **Age group (n,%)** | | | | |
| 15-19 | 33 372 (12.8%) | 5531 (3.6%) | 1757 (10.9%) | < 0.0001 |
| 20-29 | 68 006 (26.1%) | 17 979 (11.8%) | 3025 (18.8%) | |
| 30-39 | 50 752 (19.5%) | 17 817 (11.7%) | 2621 (16.3%) | |
| 40-49 | 47 302 (18.1%) | 27 494 (18.0%) | 2825 (17.5%) | |
| 50-59 | 35 868 (13.8%) | 38 819 (25.4%) | 2866 (17.8%) | |
| 60-69 | 15 522 (6.0%) | 28 505 (18.6%) | 1694 (10.5%) | |
| 70+ | 9815 (3.8%) | 16 735 (10.9%) | 1337 (8.3%) | |
| **Occupation (n,%)** | | | | |
| Non-healthcare worker | 239 173 (91.8%) | 134 054 (87.7%) | 14 551 (90.2%) | < 0.0001 |
| Healthcare worker | 21 464 (8.2%) | 18 826 (12.3%) | 1574 (9.8%) | |
| **Elderly home (n,%)** | | | | |
| No | 258 830 (99.3%) | 152 665 (99.9%) | 16 060 (99.6%) | < 0.0001 |
| Yes | 1807 (0.7%) | 215 (0.1%) | 65 (0.4%) | |
| **Origin (n,%)** | | | | |
| Danish | 198 669 (76.2%) | 136 667 (89.4%) | 13 097 (81.2%) | < 0.0001 |
| Born abroad | 47 158 (18.1%) | 14 287 (9.3%) | 2536 (15.7%) | |
| Immigrant | 14 797 (5.7%) | 1922 (1.3%) | 490 (3.0%) | |
| **Region (n,%)** | | | | |
| Hovedstaden | 108 575 (41.7%) | 56 010 (36.6%) | 6216 (38.5%) | < 0.0001 |
| Midtjylland | 50 097 (19.2%) | 30 970 (20.3%) | 3293 (20.4%) | |
| Nordjylland | 23 400 (9.0%) | 15 210 (9.9%) | 1580 (9.8%) | |
| Sjælland | 33 807 (13.0%) | 21 911 (14.3%) | 2094 (13.0%) | |
| Syddanmark | 44 522 (17.1%) | 28 691 (18.8%) | 2933 (18.2%) | |
| NA | 236 (0.1%) | 88 (0.1%) | 9 (0.1%) | |
| **Hospitalization due to COVID-19 n,%)** | | | | |
| No | 257 488 (98.8%) | 150 379 (98.4%) | 15 814 (98.1%) | < 0.0001 |
| Yes | 3149 (1.2%) | 2501 (1.6%) | 311 (1.9%) | |

Note: Charlson comorbidity scores were based on data from the Danish National Patient Register, whereas the listed comorbidities were self-reported.

**Table S3: Risk differences for post-acute symptoms reported after six, nine or twelve months, comparison of SARS-CoV-2 test-positive and test-negative participants**

| | 6 months after test (n = 22 541) | | | 9 months after test (n = 106 611) | | | 12 months after test (n = 23 728) | | |
|---|---|---|---|---|---|---|---|---|---|
| **Symptom** | **Positive (n,%)** | **Negative (n,%)** | **RD (95% CI)** | **Positive (n,%)** | **Negative (n,%)** | **RD (95% CI)** | **Positive (n,%)** | **Negative (n,%)** | **RD (95% CI)** |
| Dysosmia | 702 (9.4%) | 70 (0.5%) | 9.38 (8.66, 10.20) | 5044 (11.5%) | 422 (0.7%) | 11.56 (11.24, 11.89) | 928 (9.5%) | 112 (0.8%) | 9.26 (8.61, 9.92) |
| Dysgeusia | 587 (7.8%) | 59 (0.4%) | 7.78 (7.13, 8.44) | 4008 (9.2%) | 391 (0.6%) | 9.06 (8.76, 9.36) | 770 (7.9%) | 101 (0.7%) | 7.65 (7.05, 8.28) |
| Fatigue/exhaustion | 920 (12.3%) | 435 (2.9%) | 9.79 (8.92, 10.66) | 4905 (11.2%) | 1959 (3.1%) | 8.49 (8.14, 8.86) | 974 (9.9%) | 474 (3.4%) | 6.95 (6.24, 7.77) |
| Dyspnea | 450 (6.0%) | 122 (0.8%) | 5.70 (5.09, 6.29) | 2361 (5.4%) | 556 (0.9%) | 4.86 (4.61, 5.12) | 466 (4.8%) | 135 (1.0%) | 4.19 (3.73, 4.69) |
| Reduced strength legs/arms | 448 (6.0%) | 140 (0.9%) | 5.21 (4.57, 5.83) | 2448 (5.6%) | 715 (1.1%) | 4.71 (4.46, 5.00) | 485 (5.0%) | 169 (1.2%) | 4.04 (3.53, 4.58) |
| Sleeping legs/arms | 355 (4.7%) | 174 (1.2%) | 3.67 (3.12, 4.23) | 2053 (4.7%) | 859 (1.4%) | 3.53 (3.29, 3.78) | 433 (4.4%) | 203 (1.5%) | 3.21 (2.68, 3.74) |
| Muscle/joint pain | 429 (5.7%) | 263 (1.7%) | 4.05 (3.44, 4.73) | 2304 (5.3%) | 1236 (2.0%) | 3.41 (3.15, 3.67) | 484 (4.9%) | 273 (2.0%) | 3.17 (2.64, 3.75) |
| Headache | 502 (6.7%) | 413 (2.7%) | 4.05 (3.36, 4.79) | 2657 (6.1%) | 2003 (3.2%) | 2.93 (2.63, 3.25) | 581 (5.9%) | 452 (3.2%) | 2.69 (2.11, 3.30) |
| Dizziness | 322 (4.3%) | 210 (1.4%) | 2.86 (2.33, 3.45) | 1754 (4.0%) | 1049 (1.7%) | 2.38 (2.15, 2.61) | 354 (3.6%) | 236 (1.7%) | 1.91 (1.45, 2.42) |
| Chest pain | 234 (3.1%) | 90 (0.6%) | 2.54 (2.13, 3.00) | 1195 (2.7%) | 538 (0.9%) | 1.97 (1.79, 2.15) | 266 (2.7%) | 152 (1.1%) | 1.72 (1.34, 2.13) |
| Hot flushes/sweat | 264 (3.5%) | 225 (1.5%) | 2.04 (1.52, 2.57) | 1474 (3.4%) | 1085 (1.7%) | 1.63 (1.42, 1.85) | 309 (3.2%) | 240 (1.7%) | 1.49 (1.03, 1.99) |
| Reduced appetite | 242 (3.2%) | 153 (1.0%) | 2.08 (1.64, 2.54) | 1251 (2.9%) | 815 (1.3%) | 1.45 (1.26, 1.65) | 279 (2.8%) | 208 (1.5%) | 1.32 (0.90, 1.75) |
| Chills | 117 (1.6%) | 134 (0.9%) | 0.51 (0.18, 0.85) | 702 (1.6%) | 666 (1.1%) | 0.48 (0.33, 0.63) | 147 (1.5%) | 180 (1.3%) | 0.15 (-0.20, 0.49) |
| Red runny eyes | 92 (1.2%) | 97 (0.6%) | 0.58 (0.30, 0.91) | 583 (1.3%) | 512 (0.8%) | 0.48 (0.34, 0.61) | 147 (1.5%) | 139 (1.0%) | 0.56 (0.26, 0.92) |
| Nausea | 136 (1.8%) | 166 (1.1%) | 0.59 (0.22, 0.98) | 846 (1.9%) | 915 (1.5%) | 0.41 (0.25, 0.59) | 197 (2.0%) | 213 (1.5%) | 0.42 (0.04, 0.80) |
| Abdominal pain | 156 (2.1%) | 192 (1.3%) | 0.83 (0.41, 1.25) | 877 (2.0%) | 1005 (1.6%) | 0.35 (0.18, 0.54) | 208 (2.1%) | 213 (1.5%) | 0.51 (0.13, 0.89) |
| Diarrhoea | 119 (1.6%) | 190 (1.3%) | 0.23 (-0.11, 0.57) | 830 (1.9%) | 962 (1.5%) | 0.32 (0.14, 0.51) | 173 (1.8%) | 186 (1.3%) | 0.53 (0.17, 0.88) |
| Fever | 172 (2.3%) | 206 (1.4%) | 0.69 (0.28, 1.13) | 978 (2.2%) | 1120 (1.8%) | 0.27 (0.07, 0.47) | 212 (2.2%) | 258 (1.9%) | 0.15 (-0.24, 0.56) |
| Cough | 349 (4.6%) | 592 (3.9%) | 0.34 (-0.25, 0.96) | 2097 (4.8%) | 2789 (4.4%) | -0.02 (-0.29, 0.25) | 510 (5.2%) | 696 (5.0%) | -0.25 (-0.95, 0.36) |
| Runny nose | 267 (3.6%) | 527 (3.5%) | -0.26 (-0.80, 0.27) | 1710 (3.9%) | 2373 (3.8%) | -0.16 (-0.44, 0.09) | 399 (4.1%) | 574 (4.1%) | -0.41 (-0.96, 0.19) |
| Sore throat | 276 (3.7%) | 519 (3.5%) | -0.16 (-0.75, 0.45) | 1602 (3.7%) | 2583 (4.1%) | -0.79 (-1.04, -0.52) | 404 (4.1%) | 588 (4.2%) | -0.47 (-1.09, 0.17) |

**Table S4: Risk differences of symptoms after 6-12 months, comparing SARS-CoV-2 test-positive and test-negative participants, stratified by sex and age group**

(page 1 of 7)

| | | Female (n = 93 494) | | | Male (n = 59 386) | | |
|---|---|---|---|---|---|---|---|
| **Symptom** | **Age (years)** | **Positive (n, %)** | **Negative (n, %)** | **RD (95% CI)** | **Positive (n, %)** | **Negative (n, %)** | **RD (95% CI)** |
| Abdominal pain | 15-19 | 49 (3.2%) | 69 (3.2%) | 0.08 (-1.21, 1.45) | 9 (1.1%) | 22 (2.2%) | -1.25 (-2.54, 0.09) |
| | 20-29 | 179 (3.0%) | 198 (3.2%) | -0.09 (-0.74, 0.55) | 55 (1.8%) | 40 (1.5%) | 0.34 (-0.38, 1.08) |
| | 30-39 | 162 (3.3%) | 216 (3.3%) | 0.03 (-0.61, 0.74) | 56 (1.9%) | 51 (1.6%) | 0.34 (-0.35, 1.06) |
| | 40-49 | 180 (2.6%) | 218 (2.0%) | 0.55 (0.10, 1.02) | 69 (1.6%) | 72 (1.4%) | 0.24 (-0.27, 0.76) |
| | 50-59 | 226 (2.6%) | 251 (1.6%) | 0.99 (0.62, 1.38) | 72 (1.1%) | 70 (0.8%) | 0.29 (-0.01, 0.62) |
| | 60-69 | 98 (1.9%) | 107 (1.0%) | 0.91 (0.51, 1.30) | 44 (1.0%) | 44 (0.6%) | 0.39 (0.09, 0.69) |
| | 70+ | 23 (0.8%) | 30 (0.6%) | 0.22 (-0.13, 0.60) | 19 (0.6%) | 22 (0.4%) | 0.21 (-0.06, 0.53) |
| Dyspnea | 15-19 | 53 (3.5%) | 34 (1.6%) | 2.37 (1.09, 3.65) | 13 (1.6%) | 10 (1.0%) | 0.71 (-0.54, 1.94) |
| | 20-29 | 252 (4.3%) | 80 (1.3%) | 3.48 (2.81, 4.16) | 84 (2.7%) | 23 (0.9%) | 2.22 (1.40, 2.97) |
| | 30-39 | 305 (6.1%) | 77 (1.2%) | 5.35 (4.60, 6.10) | 123 (4.1%) | 23 (0.7%) | 3.77 (2.98, 4.63) |
| | 40-49 | 509 (7.2%) | 117 (1.1%) | 6.25 (5.61, 6.90) | 250 (5.8%) | 48 (0.9%) | 5.08 (4.37, 5.89) |
| | 50-59 | 614 (7.2%) | 171 (1.1%) | 5.94 (5.37, 6.51) | 376 (5.9%) | 71 (0.8%) | 4.98 (4.36, 5.55) |
| | 60-69 | 303 (6.0%) | 68 (0.6%) | 5.04 (4.44, 5.68) | 235 (5.3%) | 49 (0.6%) | 4.30 (3.67, 4.95) |
| | 70+ | 76 (2.7%) | 22 (0.4%) | 2.10 (1.49, 2.68) | 84 (2.8%) | 20 (0.4%) | 2.22 (1.64, 2.79) |
| Chest pain | 15-19 | 57 (3.8%) | 32 (1.5%) | 2.62 (1.43, 3.98) | 14 (1.7%) | 10 (1.0%) | 0.78 (-0.38, 2.05) |
| | 20-29 | 214 (3.6%) | 85 (1.4%) | 2.57 (1.97, 3.21) | 69 (2.2%) | 29 (1.1%) | 1.30 (0.48, 2.01) |
| | 30-39 | 182 (3.7%) | 75 (1.1%) | 2.79 (2.13, 3.50) | 85 (2.8%) | 41 (1.3%) | 1.74 (1.00, 2.52) |
| | 40-49 | 279 (4.0%) | 127 (1.2%) | 2.90 (2.36, 3.41) | 107 (2.5%) | 42 (0.8%) | 1.72 (1.19, 2.28) |
| | 50-59 | 286 (3.3%) | 133 (0.9%) | 2.46 (2.06, 2.89) | 154 (2.4%) | 59 (0.7%) | 1.64 (1.26, 2.05) |
| | 60-69 | 104 (2.1%) | 68 (0.6%) | 1.35 (0.97, 1.75) | 87 (1.9%) | 48 (0.6%) | 1.20 (0.81, 1.61) |
| | 70+ | 24 (0.8%) | 13 (0.2%) | 0.53 (0.24, 0.86) | 33 (1.1%) | 18 (0.3%) | 0.67 (0.34, 1.03) |

**Table S4: Risk differences of symptoms after 6-12 months, comparing SARS-CoV-2 test-positive and test-negative participants, stratified by sex and age group, continued**
(page 2 of 7)

| | | Female (n = 93 494) | | | Male (n = 59 386) | | |
|---|---|---|---|---|---|---|---|
| **Symptom** | **Age (years)** | **Positive (n, %)** | **Negative (n, %)** | **RD (95% CI)** | **Positive (n, %)** | **Negative (n, %)** | **RD (95% CI)** |
| Chills | 15-19 | 41 (2.7%) | 45 (2.1%) | 0.72 (-0.53, 1.85) | 6 (0.7%) | 16 (1.6%) | -0.96 (-2.04, 0.04) |
| | 20-29 | 140 (2.4%) | 127 (2.0%) | 0.41 (-0.17, 1.01) | 37 (1.2%) | 35 (1.3%) | -0.11 (-0.80, 0.53) |
| | 30-39 | 122 (2.5%) | 141 (2.1%) | 0.35 (-0.22, 0.96) | 52 (1.7%) | 59 (1.8%) | -0.07 (-0.77, 0.61) |
| | 40-49 | 147 (2.1%) | 156 (1.4%) | 0.65 (0.23, 1.08) | 51 (1.2%) | 57 (1.1%) | 0.10 (-0.34, 0.52) |
| | 50-59 | 164 (1.9%) | 173 (1.1%) | 0.78 (0.45, 1.10) | 76 (1.2%) | 53 (0.6%) | 0.54 (0.25, 0.85) |
| | 60-69 | 46 (0.9%) | 60 (0.5%) | 0.34 (0.08, 0.62) | 43 (1.0%) | 29 (0.4%) | 0.54 (0.25, 0.84) |
| | 70+ | 22 (0.8%) | 17 (0.3%) | 0.41 (0.09, 0.77) | 19 (0.6%) | 12 (0.2%) | 0.37 (0.10, 0.66) |
| Cough | 15-19 | 152 (10.1%) | 244 (11.3%) | -1.32 (-3.49, 0.80) | 55 (6.6%) | 100 (9.8%) | -3.49 (-6.02, -0.92) |
| | 20-29 | 399 (6.8%) | 540 (8.6%) | -1.86 (-2.85, -0.89) | 181 (5.8%) | 180 (6.7%) | -0.92 (-2.22, 0.42) |
| | 30-39 | 273 (5.5%) | 387 (5.9%) | -0.36 (-1.21, 0.49) | 169 (5.6%) | 171 (5.2%) | 0.41 (-0.73, 1.58) |
| | 40-49 | 363 (5.2%) | 590 (5.5%) | -0.33 (-1.00, 0.34) | 171 (3.9%) | 247 (4.6%) | -0.73 (-1.57, 0.14) |
| | 50-59 | 441 (5.2%) | 642 (4.2%) | 0.98 (0.41, 1.54) | 261 (4.1%) | 277 (3.3%) | 0.76 (0.16, 1.37) |
| | 60-69 | 204 (4.0%) | 316 (2.8%) | 1.15 (0.52, 1.74) | 117 (2.6%) | 196 (2.5%) | 0.10 (-0.43, 0.68) |
| | 70+ | 89 (3.2%) | 102 (1.9%) | 1.19 (0.45, 1.92) | 81 (2.7%) | 85 (1.5%) | 1.15 (0.52, 1.79) |
| Diarrhoea | 15-19 | 31 (2.1%) | 40 (1.8%) | 0.25 (-0.79, 1.32) | 13 (1.6%) | 19 (1.9%) | -0.36 (-1.75, 0.90) |
| | 20-29 | 136 (2.3%) | 158 (2.5%) | -0.19 (-0.84, 0.40) | 55 (1.8%) | 44 (1.6%) | 0.18 (-0.57, 0.91) |
| | 30-39 | 147 (3.0%) | 183 (2.8%) | 0.22 (-0.48, 0.85) | 69 (2.3%) | 48 (1.5%) | 0.91 (0.15, 1.64) |
| | 40-49 | 166 (2.4%) | 177 (1.6%) | 0.73 (0.31, 1.20) | 65 (1.5%) | 89 (1.7%) | -0.19 (-0.71, 0.31) |
| | 50-59 | 199 (2.3%) | 218 (1.4%) | 0.89 (0.53, 1.28) | 80 (1.2%) | 82 (1.0%) | 0.27 (-0.08, 0.60) |
| | 60-69 | 74 (1.5%) | 128 (1.1%) | 0.28 (-0.05, 0.64) | 43 (1.0%) | 75 (1.0%) | 0.00 (-0.30, 0.32) |
| | 70+ | 25 (0.9%) | 39 (0.7%) | 0.13 (-0.23, 0.53) | 19 (0.6%) | 38 (0.7%) | -0.04 (-0.35, 0.29) |

**Table S4: Risk differences of symptoms after 6-12 months, comparing SARS-CoV-2 test-positive and test-negative participants, stratified by sex and age group, continued**
(page 3 of 7)

| | | Female (n = 93 494) | | | Male (n = 59 386) | | |
|---|---|---|---|---|---|---|---|
| **Symptom** | **Age (years)** | **Positive (n, %)** | **Negative (n, %)** | **RD (95% CI)** | **Positive (n, %)** | **Negative (n, %)** | **RD (95% CI)** |
| Dizziness | 15-19 | 84 (5.6%) | 75 (3.5%) | 2.38 (0.84, 3.99) | 12 (1.4%) | 24 (2.4%) | -1.03 (-2.48, 0.30) |
| | 20-29 | 305 (5.2%) | 218 (3.5%) | 1.92 (1.09, 2.69) | 70 (2.2%) | 55 (2.1%) | 0.25 (-0.61, 1.00) |
| | 30-39 | 321 (6.5%) | 197 (3.0%) | 3.74 (2.83, 4.61) | 98 (3.2%) | 55 (1.7%) | 1.70 (0.87, 2.55) |
| | 40-49 | 400 (5.7%) | 226 (2.1%) | 3.68 (3.05, 4.29) | 140 (3.2%) | 65 (1.2%) | 2.05 (1.42, 2.67) |
| | 50-59 | 438 (5.1%) | 260 (1.7%) | 3.42 (2.87, 3.94) | 160 (2.5%) | 79 (0.9%) | 1.53 (1.11, 1.94) |
| | 60-69 | 164 (3.2%) | 119 (1.1%) | 2.06 (1.56, 2.58) | 108 (2.4%) | 52 (0.7%) | 1.62 (1.17, 2.07) |
| | 70+ | 64 (2.3%) | 33 (0.6%) | 1.51 (0.98, 2.05) | 66 (2.2%) | 37 (0.7%) | 1.38 (0.88, 1.90) |
| Fatigue/exhaustion | 15-19 | 179 (11.8%) | 108 (5.0%) | 7.37 (5.41, 9.49) | 53 (6.3%) | 46 (4.5%) | 1.98 (-0.32, 4.25) |
| | 20-29 | 651 (11.1%) | 339 (5.4%) | 5.99 (5.00, 7.02) | 215 (6.9%) | 125 (4.7%) | 2.41 (1.13, 3.72) |
| | 30-39 | 764 (15.4%) | 310 (4.7%) | 10.93 (9.74, 12.11) | 277 (9.2%) | 123 (3.8%) | 5.61 (4.37, 6.85) |
| | 40-49 | 1061 (15.1%) | 459 (4.3%) | 10.84 (9.94, 11.79) | 510 (11.8%) | 208 (3.9%) | 7.93 (6.84, 9.12) |
| | 50-59 | 1218 (14.3%) | 501 (3.2%) | 10.91 (10.11, 11.74) | 692 (10.8%) | 216 (2.6%) | 8.17 (7.28, 8.92) |
| | 60-69 | 525 (10.4%) | 212 (1.9%) | 8.29 (7.47, 9.15) | 359 (8.0%) | 118 (1.5%) | 6.35 (5.56, 7.19) |
| | 70+ | 149 (5.3%) | 52 (1.0%) | 4.24 (3.41, 5.09) | 146 (4.9%) | 51 (0.9%) | 3.92 (3.10, 4.75) |
| Fever | 15-19 | 59 (3.9%) | 98 (4.5%) | -0.66 (-2.01, 0.83) | 24 (2.9%) | 31 (3.0%) | -0.19 (-1.86, 1.53) |
| | 20-29 | 225 (3.8%) | 240 (3.8%) | 0.03 (-0.72, 0.73) | 75 (2.4%) | 78 (2.9%) | -0.51 (-1.37, 0.31) |
| | 30-39 | 157 (3.2%) | 200 (3.0%) | 0.14 (-0.49, 0.82) | 90 (3.0%) | 73 (2.2%) | 0.82 (-0.01, 1.66) |
| | 40-49 | 169 (2.4%) | 224 (2.1%) | 0.32 (-0.18, 0.77) | 87 (2.0%) | 94 (1.8%) | 0.24 (-0.32, 0.78) |
| | 50-59 | 196 (2.3%) | 259 (1.7%) | 0.60 (0.23, 0.98) | 110 (1.7%) | 90 (1.1%) | 0.64 (0.25, 1.02) |
| | 60-69 | 60 (1.2%) | 111 (1.0%) | 0.17 (-0.16, 0.50) | 57 (1.3%) | 53 (0.7%) | 0.56 (0.24, 0.92) |
| | 70+ | 22 (0.8%) | 19 (0.4%) | 0.39 (0.06, 0.77) | 31 (1.0%) | 14 (0.2%) | 0.72 (0.39, 1.08) |

**Table S4: Risk differences of symptoms after 6-12 months, comparing SARS-CoV-2 test-positive and test-negative participants, stratified by sex and age group, continued**
(page 4 of 7)

| | | Female (n = 93 494) | | | Male (n = 59 386) | | |
|---|---|---|---|---|---|---|---|
| **Symptom** | **Age (years)** | **Positive (n, %)** | **Negative (n, %)** | **RD (95% CI)** | **Positive (n, %)** | **Negative (n, %)** | **RD (95% CI)** |
| Headache | 15-19 | 121 (8.0%) | 120 (5.5%) | 2.59 (0.99, 4.33) | 34 (4.1%) | 51 (5.0%) | -0.99 (-2.96, 0.86) |
| | 20-29 | 460 (7.8%) | 329 (5.2%) | 2.68 (1.80, 3.63) | 163 (5.2%) | 129 (4.8%) | 0.44 (-0.70, 1.59) |
| | 30-39 | 504 (10.2%) | 326 (5.0%) | 5.26 (4.28, 6.17) | 206 (6.8%) | 135 (4.1%) | 2.77 (1.53, 3.92) |
| | 40-49 | 599 (8.5%) | 421 (3.9%) | 4.59 (3.86, 5.33) | 265 (6.1%) | 200 (3.8%) | 2.37 (1.49, 3.28) |
| | 50-59 | 657 (7.7%) | 482 (3.1%) | 4.52 (3.90, 5.16) | 244 (3.8%) | 219 (2.6%) | 1.21 (0.62, 1.82) |
| | 60-69 | 241 (4.8%) | 223 (2.0%) | 2.71 (2.07, 3.39) | 146 (3.3%) | 138 (1.8%) | 1.48 (0.91, 2.10) |
| | 70+ | 59 (2.1%) | 58 (1.1%) | 0.99 (0.40, 1.59) | 41 (1.4%) | 37 (0.7%) | 0.71 (0.24, 1.19) |
| Hot flushes/sweat | 15-19 | 66 (4.4%) | 71 (3.3%) | 1.24 (-0.13, 2.66) | 17 (2.0%) | 21 (2.1%) | -0.02 (-1.41, 1.52) |
| | 20-29 | 253 (4.3%) | 229 (3.6%) | 0.75 (0.01, 1.47) | 63 (2.0%) | 49 (1.8%) | 0.24 (-0.53, 1.01) |
| | 30-39 | 227 (4.6%) | 188 (2.9%) | 1.83 (1.15, 2.57) | 90 (3.0%) | 71 (2.2%) | 0.89 (0.06, 1.73) |
| | 40-49 | 358 (5.1%) | 261 (2.4%) | 2.70 (2.13, 3.29) | 118 (2.7%) | 98 (1.8%) | 0.90 (0.28, 1.53) |
| | 50-59 | 386 (4.5%) | 228 (1.5%) | 2.98 (2.53, 3.47) | 161 (2.5%) | 98 (1.2%) | 1.33 (0.89, 1.75) |
| | 60-69 | 152 (3.0%) | 120 (1.1%) | 1.83 (1.34, 2.34) | 74 (1.7%) | 67 (0.9%) | 0.75 (0.36, 1.14) |
| | 70+ | 48 (1.7%) | 29 (0.5%) | 1.08 (0.63, 1.59) | 34 (1.1%) | 20 (0.4%) | 0.73 (0.34, 1.13) |
| Dysosmia | 15-19 | 207 (13.7%) | 52 (2.4%) | 11.77 (9.80, 13.72) | 83 (9.9%) | 18 (1.8%) | 8.46 (6.08, 10.72) |
| | 20-29 | 854 (14.5%) | 77 (1.2%) | 13.60 (12.61, 14.61) | 401 (12.8%) | 17 (0.6%) | 12.57 (11.3, 13.82) |
| | 30-39 | 743 (15.0%) | 64 (1.0%) | 14.13 (13.07, 15.19) | 370 (12.3%) | 23 (0.7%) | 11.84 (10.65, 13.13) |
| | 40-49 | 1057 (15.0%) | 106 (1.0%) | 14.01 (13.14, 14.90) | 484 (11.2%) | 38 (0.7%) | 10.55 (9.62, 11.62) |
| | 50-59 | 1068 (12.5%) | 92 (0.6%) | 11.79 (11.07, 12.49) | 531 (8.3%) | 40 (0.5%) | 7.80 (7.11, 8.47) |
| | 60-69 | 407 (8.0%) | 39 (0.3%) | 7.54 (6.78, 8.31) | 269 (6.0%) | 19 (0.2%) | 5.69 (4.95, 6.38) |
| | 70+ | 109 (3.9%) | 9 (0.2%) | 3.66 (2.99, 4.42) | 91 (3.1%) | 10 (0.2%) | 2.85 (2.27, 3.49) |

**Table S4: Risk differences of symptoms after 6-12 months, comparing SARS-CoV-2 test-positive and test-negative participants, stratified by sex and age group, continued**
(page 5 of 7)

| | | Female (n = 93 494) | | | Male (n = 59 386) | | |
|---|---|---|---|---|---|---|---|
| **Symptom** | **Age (years)** | **Positive (n, %)** | **Negative (n, %)** | **RD (95% CI)** | **Positive (n, %)** | **Negative (n, %)** | **RD (95% CI)** |
| Dysgeusia | 15-19 | 155 (10.3%) | 27 (1.2%) | 9.56 (7.87, 11.23) | 59 (7.0%) | 5 (0.5%) | 6.97 (5.25, 8.92) |
| | 20-29 | 635 (10.8%) | 64 (1.0%) | 10.19 (9.30, 11.09) | 303 (9.7%) | 18 (0.7%) | 9.54 (8.38, 10.74) |
| | 30-39 | 591 (11.9%) | 63 (1.0%) | 11.21 (10.26, 12.31) | 273 (9.0%) | 19 (0.6%) | 8.85 (7.68, 10.01) |
| | 40-49 | 858 (12.2%) | 107 (1.0%) | 11.26 (10.51, 11.97) | 378 (8.7%) | 34 (0.6%) | 8.25 (7.39, 9.16) |
| | 50-59 | 917 (10.7%) | 88 (0.6%) | 10.05 (9.42, 10.75) | 428 (6.7%) | 37 (0.4%) | 6.24 (5.65, 6.88) |
| | 60-69 | 348 (6.9%) | 46 (0.4%) | 6.26 (5.57, 6.96) | 221 (5.0%) | 20 (0.3%) | 4.56 (3.93, 5.22) |
| | 70+ | 114 (4.0%) | 15 (0.3%) | 3.60 (2.90, 4.32) | 85 (2.9%) | 8 (0.1%) | 2.58 (2.03, 3.20) |
| Muscle/joint pain | 15-19 | 64 (4.2%) | 74 (3.4%) | 0.89 (-0.42, 2.23) | 15 (1.8%) | 14 (1.4%) | 0.46 (-0.71, 1.64) |
| | 20-29 | 293 (5.0%) | 218 (3.5%) | 1.61 (0.86, 2.35) | 96 (3.1%) | 68 (2.5%) | 0.59 (-0.27, 1.53) |
| | 30-39 | 363 (7.3%) | 238 (3.6%) | 3.79 (2.91, 4.61) | 150 (5.0%) | 99 (3.0%) | 2.02 (0.97, 3.08) |
| | 40-49 | 525 (7.5%) | 291 (2.7%) | 4.76 (4.08, 5.39) | 217 (5.0%) | 115 (2.2%) | 2.88 (2.14, 3.64) |
| | 50-59 | 606 (7.1%) | 266 (1.7%) | 5.30 (4.72, 5.92) | 331 (5.2%) | 123 (1.5%) | 3.68 (3.11, 4.29) |
| | 60-69 | 241 (4.8%) | 113 (1.0%) | 3.66 (3.04, 4.29) | 180 (4.0%) | 87 (1.1%) | 2.84 (2.27, 3.44) |
| | 70+ | 63 (2.2%) | 29 (0.5%) | 1.66 (1.08, 2.28) | 73 (2.5%) | 37 (0.7%) | 1.74 (1.22, 2.31) |
| Nausea | 15-19 | 57 (3.8%) | 70 (3.2%) | 0.66 (-0.75, 2.09) | 10 (1.2%) | 19 (1.9%) | -0.80 (-2.11, 0.45) |
| | 20-29 | 223 (3.8%) | 216 (3.4%) | 0.44 (-0.31, 1.23) | 45 (1.4%) | 33 (1.2%) | 0.28 (-0.43, 0.99) |
| | 30-39 | 163 (3.3%) | 214 (3.3%) | 0.08 (-0.69, 0.85) | 52 (1.7%) | 58 (1.8%) | -0.03 (-0.76, 0.68) |
| | 40-49 | 178 (2.5%) | 197 (1.8%) | 0.72 (0.25, 1.19) | 42 (1.0%) | 49 (0.9%) | 0.04 (-0.35, 0.49) |
| | 50-59 | 203 (2.4%) | 214 (1.4%) | 0.96 (0.61, 1.35) | 59 (0.9%) | 44 (0.5%) | 0.39 (0.10, 0.65) |
| | 60-69 | 72 (1.4%) | 110 (1.0%) | 0.39 (0.03, 0.77) | 30 (0.7%) | 29 (0.4%) | 0.27 (0.02, 0.51) |
| | 70+ | 31 (1.1%) | 25 (0.5%) | 0.55 (0.18, 0.94) | 14 (0.5%) | 16 (0.3%) | 0.15 (-0.08, 0.38) |

**Table S4: Risk differences of symptoms after 6-12 months, comparing SARS-CoV-2 test-positive and test-negative participants, stratified by sex and age group, continued**
(page 6 of 7)

| | | Female (n = 93 494) | | | Male (n = 59 386) | | |
|---|---|---|---|---|---|---|---|
| **Symptom** | **Age (years)** | **Positive (n, %)** | **Negative (n, %)** | **RD (95% CI)** | **Positive (n, %)** | **Negative (n, %)** | **RD (95% CI)** |
| Red runny eyes | 15-19 | 27 (1.8%) | 54 (2.5%) | -0.78 (-1.78, 0.22) | 6 (0.7%) | 19 (1.9%) | -1.26 (-2.38, -0.16) |
| | 20-29 | 94 (1.6%) | 92 (1.5%) | 0.15 (-0.32, 0.63) | 35 (1.1%) | 20 (0.7%) | 0.42 (-0.12, 0.94) |
| | 30-39 | 88 (1.8%) | 82 (1.2%) | 0.57 (0.08, 1.10) | 40 (1.3%) | 27 (0.8%) | 0.55 (0.02, 1.09) |
| | 40-49 | 137 (1.9%) | 113 (1.0%) | 0.92 (0.54, 1.32) | 39 (0.9%) | 31 (0.6%) | 0.31 (-0.05, 0.67) |
| | 50-59 | 155 (1.8%) | 126 (0.8%) | 0.98 (0.65, 1.28) | 63 (1.0%) | 57 (0.7%) | 0.29 (0.00, 0.58) |
| | 60-69 | 59 (1.2%) | 53 (0.5%) | 0.65 (0.32, 0.95) | 38 (0.9%) | 41 (0.5%) | 0.30 (0.01, 0.59) |
| | 70+ | 20 (0.7%) | 15 (0.3%) | 0.38 (0.09, 0.70) | 21 (0.7%) | 18 (0.3%) | 0.33 (0.05, 0.66) |
| Reduced appetite | 15-19 | 115 (7.6%) | 71 (3.3%) | 4.89 (3.24, 6.62) | 32 (3.8%) | 15 (1.5%) | 2.63 (1.10, 4.28) |
| | 20-29 | 297 (5.0%) | 200 (3.2%) | 2.08 (1.30, 2.81) | 89 (2.8%) | 37 (1.4%) | 1.68 (0.91, 2.43) |
| | 30-39 | 215 (4.3%) | 173 (2.6%) | 1.86 (1.11, 2.61) | 73 (2.4%) | 43 (1.3%) | 1.24 (0.51, 2.01) |
| | 40-49 | 222 (3.2%) | 181 (1.7%) | 1.53 (1.03, 2.04) | 86 (2.0%) | 53 (1.0%) | 1.03 (0.52, 1.56) |
| | 50-59 | 239 (2.8%) | 164 (1.1%) | 1.72 (1.35, 2.11) | 116 (1.8%) | 53 (0.6%) | 1.17 (0.79, 1.52) |
| | 60-69 | 101 (2.0%) | 90 (0.8%) | 1.11 (0.73, 1.54) | 62 (1.4%) | 35 (0.4%) | 0.86 (0.54, 1.21) |
| | 70+ | 70 (2.5%) | 33 (0.6%) | 1.63 (1.10, 2.17) | 55 (1.8%) | 28 (0.5%) | 1.14 (0.73, 1.64) |
| Reduced strength legs/arms | 15-19 | 50 (3.3%) | 39 (1.8%) | 1.75 (0.54, 2.95) | 17 (2.0%) | 5 (0.5%) | 1.76 (0.61, 3.03) |
| | 20-29 | 195 (3.3%) | 84 (1.3%) | 2.25 (1.64, 2.85) | 84 (2.7%) | 21 (0.8%) | 2.17 (1.47, 2.92) |
| | 30-39 | 294 (5.9%) | 112 (1.7%) | 4.63 (3.81, 5.41) | 140 (4.6%) | 48 (1.5%) | 3.43 (2.58, 4.36) |
| | 40-49 | 488 (6.9%) | 147 (1.4%) | 5.79 (5.16, 6.43) | 237 (5.5%) | 79 (1.5%) | 4.03 (3.27, 4.80) |
| | 50-59 | 673 (7.9%) | 195 (1.3%) | 6.63 (6.03, 7.25) | 421 (6.6%) | 86 (1.0%) | 5.34 (4.67, 5.95) |
| | 60-69 | 315 (6.2%) | 75 (0.7%) | 5.32 (4.68, 6.02) | 234 (5.2%) | 56 (0.7%) | 4.09 (3.51, 4.68) |
| | 70+ | 104 (3.7%) | 36 (0.7%) | 2.71 (2.03, 3.38) | 129 (4.3%) | 41 (0.7%) | 3.11 (2.42, 3.83) |

**Table S4: Risk differences of symptoms after 6-12 months, comparing SARS-CoV-2 test-positive and test-negative participants, stratified by sex and age group, continued**
(page 7 of 7)

| | | Female (n = 93 494) | | | Male (n = 59 386) | | |
|---|---|---|---|---|---|---|---|
| **Symptom** | **Age (years)** | **Positive (n, %)** | **Negative (n, %)** | **RD (95% CI)** | **Positive (n, %)** | **Negative (n, %)** | **RD (95% CI)** |
| Runny nose | 15-19 | 122 (8.1%) | 206 (9.5%) | -1.52 (-3.39, 0.43) | 41 (4.9%) | 87 (8.5%) | -3.82 (-6.18, -1.53) |
| | 20-29 | 368 (6.3%) | 478 (7.6%) | -1.39 (-2.29, -0.45) | 145 (4.6%) | 148 (5.5%) | -0.89 (-2.08, 0.36) |
| | 30-39 | 250 (5.0%) | 389 (5.9%) | -0.89 (-1.73, -0.02) | 122 (4.0%) | 140 (4.3%) | -0.22 (-1.21, 0.77) |
| | 40-49 | 285 (4.0%) | 504 (4.7%) | -0.66 (-1.23, -0.06) | 157 (3.6%) | 217 (4.1%) | -0.47 (-1.20, 0.32) |
| | 50-59 | 348 (4.1%) | 536 (3.5%) | 0.58 (0.04, 1.06) | 175 (2.7%) | 213 (2.5%) | 0.19 (-0.30, 0.75) |
| | 60-69 | 157 (3.1%) | 249 (2.2%) | 0.83 (0.37, 1.40) | 107 (2.4%) | 151 (1.9%) | 0.44 (-0.11, 0.97) |
| | 70+ | 51 (1.8%) | 71 (1.3%) | 0.44 (-0.14, 1.03) | 48 (1.6%) | 85 (1.5%) | 0.09 (-0.41, 0.65) |
| Sleeping legs/arms | 15-19 | 54 (3.6%) | 41 (1.9%) | 1.91 (0.68, 3.14) | 12 (1.4%) | 10 (1.0%) | 0.51 (-0.61, 1.60) |
| | 20-29 | 193 (3.3%) | 123 (2.0%) | 1.49 (0.88, 2.12) | 71 (2.3%) | 38 (1.4%) | 0.99 (0.25, 1.82) |
| | 30-39 | 301 (6.1%) | 147 (2.2%) | 4.11 (3.34, 4.90) | 122 (4.0%) | 45 (1.4%) | 2.90 (2.05, 3.82) |
| | 40-49 | 475 (6.7%) | 201 (1.9%) | 4.99 (4.39, 5.66) | 169 (3.9%) | 72 (1.4%) | 2.60 (1.91, 3.33) |
| | 50-59 | 557 (6.5%) | 228 (1.5%) | 4.98 (4.37, 5.54) | 297 (4.6%) | 81 (1.0%) | 3.59 (3.05, 4.14) |
| | 60-69 | 255 (5.0%) | 109 (1.0%) | 3.85 (3.26, 4.41) | 167 (3.7%) | 68 (0.9%) | 2.65 (2.14, 3.22) |
| | 70+ | 81 (2.9%) | 35 (0.7%) | 2.02 (1.43, 2.65) | 87 (2.9%) | 38 (0.7%) | 2.01 (1.40, 2.58) |
| Sore throat | 15-19 | 128 (8.5%) | 223 (10.3%) | -1.96 (-3.95, -0.02) | 41 (4.9%) | 97 (9.5%) | -4.93 (-7.47, -2.54) |
| | 20-29 | 384 (6.5%) | 518 (8.3%) | -1.77 (-2.77, -0.78) | 146 (4.7%) | 156 (5.8%) | -1.16 (-2.26, 0.12) |
| | 30-39 | 242 (4.9%) | 387 (5.9%) | -1.00 (-1.78, -0.17) | 112 (3.7%) | 145 (4.4%) | -0.72 (-1.70, 0.22) |
| | 40-49 | 324 (4.6%) | 613 (5.7%) | -1.10 (-1.75, -0.43) | 135 (3.1%) | 195 (3.7%) | -0.57 (-1.25, 0.17) |
| | 50-59 | 341 (4.0%) | 608 (3.9%) | 0.04 (-0.46, 0.56) | 146 (2.3%) | 192 (2.3%) | -0.01 (-0.51, 0.45) |
| | 60-69 | 138 (2.7%) | 296 (2.7%) | 0.05 (-0.46, 0.57) | 65 (1.5%) | 130 (1.7%) | -0.20 (-0.63, 0.22) |
| | 70+ | 43 (1.5%) | 80 (1.5%) | 0.00 (-0.48, 0.55) | 37 (1.2%) | 50 (0.9%) | 0.33 (-0.13, 0.79) |

**Table S5: Risk differences of health problems after 6-12 months, comparing SARS-CoV-2 test-positive and test-negative participants, stratified by sex and age group**
(page 1 of 2)

| | | Female (n = 93 494) | | | Male (n = 59 386) | | |
|---|---|---|---|---|---|---|---|
| **Medical diagnosis** | **Age (years)** | **Positive (n, %)** | **Negative (n, %)** | **RD (95% CI)** | **Positive (n, %)** | **Negative (n, %)** | **RD (95% CI)** |
| Difficulties concentrating | 15-19 | 407 (33.0%) | 161 (9.6%) | 25.16 (22.03, 28.21) | 123 (17.1%) | 43 (5.1%) | 13.32 (10.04, 16.98) |
| | 20-29 | 1712 (33.8%) | 279 (5.4%) | 30.19 (28.87, 31.74) | 628 (22.5%) | 102 (4.5%) | 19.83 (17.91, 21.70) |
| | 30-39 | 1711 (37.9%) | 316 (5.5%) | 33.70 (32.16, 35.29) | 754 (27.0%) | 100 (3.4%) | 25.20 (23.36, 27.03) |
| | 40-49 | 2562 (39.5%) | 442 (4.7%) | 35.55 (34.32, 36.79) | 1136 (27.8%) | 156 (3.2%) | 25.43 (23.97, 26.91) |
| | 50-59 | 2966 (37.6%) | 531 (3.8%) | 33.68 (32.49, 34.80) | 1638 (26.8%) | 206 (2.6%) | 24.24 (23.15, 25.40) |
| | 60-69 | 1324 (27.6%) | 197 (1.9%) | 24.85 (23.54, 26.11) | 926 (21.6%) | 130 (1.7%) | 19.02 (17.79, 20.22) |
| | 70+ | 428 (15.7%) | 61 (1.2%) | 13.58 (12.20, 14.98) | 405 (14.2%) | 88 (1.6%) | 11.46 (10.18, 12.67) |
| Memory issues | 15-19 | 350 (26.0%) | 103 (5.5%) | 22.46 (19.68, 25.17) | 105 (13.7%) | 34 (3.6%) | 11.14 (7.96, 14.23) |
| | 20-29 | 1502 (28.1%) | 286 (5.3%) | 24.58 (23.12, 25.99) | 496 (17.0%) | 91 (3.8%) | 14.75 (12.99, 16.45) |
| | 30-39 | 1616 (36.1%) | 331 (5.9%) | 31.76 (30.22, 33.35) | 592 (21.0%) | 93 (3.1%) | 19.24 (17.51, 21.05) |
| | 40-49 | 2532 (40.0%) | 481 (5.2%) | 35.72 (34.34, 37.03) | 1072 (26.3%) | 175 (3.6%) | 23.54 (22.17, 25.09) |
| | 50-59 | 3075 (39.4%) | 583 (4.3%) | 35.20 (34.08, 36.46) | 1582 (26.0%) | 245 (3.1%) | 22.84 (21.70, 24.04) |
| | 60-69 | 1376 (29.0%) | 260 (2.5%) | 25.65 (24.41, 26.87) | 944 (22.5%) | 160 (2.2%) | 19.27 (18.09, 20.50) |
| | 70+ | 450 (17.0%) | 82 (1.7%) | 14.18 (12.84, 15.56) | 457 (16.6%) | 133 (2.6%) | 12.62 (11.32, 14.00) |
| Mental exhaustion | 15-19 | 537 (46.8%) | 280 (18.9%) | 29.19 (25.48, 33.05) | 176 (24.3%) | 82 (9.8%) | 15.76 (11.94, 19.56) |
| | 20-29 | 2228 (46.0%) | 634 (14.2%) | 33.14 (31.27, 34.90) | 796 (28.8%) | 251 (11.7%) | 18.53 (16.27, 20.63) |
| | 30-39 | 2047 (47.6%) | 638 (12.5%) | 36.17 (34.33, 38.04) | 941 (34.7%) | 227 (8.4%) | 27.70 (25.61, 29.80) |
| | 40-49 | 3049 (48.9%) | 920 (10.4%) | 39.11 (37.73, 40.48) | 1421 (35.7%) | 346 (7.5%) | 28.97 (27.13, 30.68) |
| | 50-59 | 3652 (47.0%) | 1069 (8.1%) | 38.96 (37.76, 40.15) | 2089 (34.3%) | 457 (6.0%) | 28.44 (27.12, 29.92) |
| | 60-69 | 1737 (36.3%) | 474 (4.7%) | 30.96 (29.54, 32.37) | 1145 (26.8%) | 273 (3.7%) | 22.30 (21.01, 23.62) |
| | 70+ | 544 (20.0%) | 98 (2.0%) | 17.36 (15.78, 18.81) | 448 (15.6%) | 128 (2.4%) | 12.27 (10.92, 13.54) |

**Table S5: Risk differences of health problems after 6-12 months, comparing SARS-CoV-2 test-positive and test-negative participants, stratified by sex and age group, continued**
(page 2 of 2)

| | | Female (n = 93 494) | | | Male (n = 59 386) | | |
|---|---|---|---|---|---|---|---|
| **Medical diagnosis** | **Age (years)** | **Positive (n, %)** | **Negative (n, %)** | **RD (95% CI)** | **Positive (n, %)** | **Negative (n, %)** | **RD (95% CI)** |
| Physical exhaustion | 15-19 | 608 (46.4%) | 213 (12.2%) | 36.32 (33.11, 39.50) | 179 (23.2%) | 58 (6.5%) | 18.47 (14.70, 22.24) |
| | 20-29 | 2466 (47.3%) | 547 (10.6%) | 38.42 (36.69, 40.07) | 895 (31.0%) | 195 (8.4%) | 24.67 (22.43, 26.79) |
| | 30-39 | 2290 (51.4%) | 593 (10.9%) | 41.81 (40.00, 43.65) | 1121 (40.3%) | 205 (7.1%) | 34.97 (32.87, 37.18) |
| | 40-49 | 3544 (55.3%) | 875 (9.5%) | 46.54 (45.16, 47.81) | 1798 (44.4%) | 330 (6.9%) | 38.63 (36.89, 40.32) |
| | 50-59 | 4302 (54.7%) | 1109 (8.2%) | 46.74 (45.52, 48.02) | 2639 (43.5%) | 446 (5.8%) | 37.94 (36.57, 39.36) |
| | 60-69 | 2262 (47.6%) | 580 (5.8%) | 41.15 (39.79, 42.54) | 1663 (39.7%) | 335 (4.7%) | 34.05 (32.45, 35.43) |
| | 70+ | 887 (34.1%) | 176 (3.7%) | 29.34 (27.48, 31.20) | 838 (30.5%) | 217 (4.2%) | 24.64 (22.85, 26.40) |
| Sleep problems | 15-19 | 292 (23.9%) | 193 (11.3%) | 13.45 (10.61, 16.43) | 112 (15.6%) | 60 (7.1%) | 9.14 (5.73, 12.53) |
| | 20-29 | 1054 (21.6%) | 425 (8.7%) | 13.72 (12.13, 15.22) | 431 (15.9%) | 168 (7.7%) | 9.02 (7.07, 10.90) |
| | 30-39 | 988 (23.2%) | 458 (8.6%) | 15.30 (13.77, 16.83) | 466 (17.6%) | 157 (5.7%) | 12.72 (10.91, 14.43) |
| | 40-49 | 1600 (27.1%) | 740 (8.4%) | 19.03 (17.79, 20.41) | 774 (20.4%) | 292 (6.4%) | 14.31 (12.81, 15.78) |
| | 50-59 | 2278 (33.4%) | 1006 (8.3%) | 23.98 (23.81, 26.14) | 1214 (21.3%) | 380 (5.2%) | 16.02 (14.82, 17.20) |
| | 60-69 | 1071 (26.0%) | 483 (5.4%) | 19.96 (18.57, 21.30) | 780 (19.4%) | 257 (3.7%) | 14.92 (13.73, 16.13) |
| | 70+ | 443 (19.1%) | 157 (3.6%) | 14.52 (13.14, 16.11) | 347 (12.8%) | 160 (3.1%) | 8.82 (7.57, 10.03) |

**Table S6: Risk differences of self-reported diagnoses after 6-12 months, comparing SARS-CoV-2 test-positive and test-negative participants, stratified by sex and age group**
(page 1 of 2)

| | | Female (n = 93 494) | | | Male (n = 59 386) | | |
|---|---|---|---|---|---|---|---|
| **Medical diagnosis** | **Age (years)** | **Positive (n, %)** | **Negative (n, %)** | **RD (95% CI)** | **Positive (n, %)** | **Negative (n, %)** | **RD (95% CI)** |
| Depression | 15-19 | 87 (6.2%) | 99 (5.1%) | 1.16 (-0.47, 2.78) | 29 (3.6%) | 26 (2.7%) | 0.94 (-0.77, 2.70) |
| | 20-29 | 244 (4.8%) | 177 (3.3%) | 1.62 (0.81, 2.50) | 105 (3.6%) | 82 (3.4%) | 0.29 (-0.81, 1.36) |
| | 30-39 | 188 (4.5%) | 118 (2.2%) | 2.51 (1.73, 3.32) | 91 (3.3%) | 61 (2.1%) | 1.33 (0.38, 2.26) |
| | 40-49 | 194 (3.2%) | 182 (2.0%) | 1.26 (0.69, 1.82) | 119 (3.0%) | 92 (1.9%) | 1.10 (0.37, 1.86) |
| | 50-59 | 227 (3.0%) | 284 (2.1%) | 0.87 (0.43, 1.30) | 174 (2.9%) | 180 (2.3%) | 0.57 (0.02, 1.14) |
| | 60-69 | 116 (2.6%) | 200 (2.0%) | 0.48 (-0.05, 0.97) | 119 (2.9%) | 158 (2.2%) | 0.62 (0.11, 1.17) |
| | 70+ | 101 (3.9%) | 112 (2.3%) | 1.38 (0.66, 2.11) | 89 (3.2%) | 99 (1.8%) | 1.08 (0.46, 1.68) |
| Anxiety | 15-19 | 77 (5.9%) | 103 (5.6%) | 0.26 (-1.48, 1.97) | 24 (3.0%) | 18 (1.9%) | 1.17 (-0.31, 2.79) |
| | 20-29 | 310 (6.2%) | 174 (3.3%) | 3.10 (2.21, 3.98) | 114 (3.9%) | 70 (2.8%) | 1.16 (0.16, 2.23) |
| | 30-39 | 218 (5.0%) | 126 (2.2%) | 2.94 (2.08, 3.75) | 84 (3.0%) | 48 (1.6%) | 1.48 (0.59, 2.25) |
| | 40-49 | 211 (3.3%) | 178 (1.8%) | 1.53 (1.01, 2.06) | 104 (2.5%) | 79 (1.6%) | 0.98 (0.40, 1.54) |
| | 50-59 | 237 (3.0%) | 274 (1.9%) | 1.03 (0.61, 1.46) | 150 (2.4%) | 154 (1.9%) | 0.51 (0.04, 0.96) |
| | 60-69 | 109 (2.3%) | 213 (2.0%) | 0.19 (-0.28, 0.67) | 99 (2.3%) | 158 (2.1%) | 0.17 (-0.35, 0.64) |
| | 70+ | 88 (3.2%) | 104 (2.1%) | 1.04 (0.38, 1.69) | 75 (2.6%) | 84 (1.5%) | 0.90 (0.32, 1.50) |
| PTSD | 15-19 | 19 (1.3%) | 19 (0.9%) | 0.39 (-0.29, 1.16) | 6 (0.7%) | 7 (0.7%) | 0.03 (-0.76, 0.79) |
| | 20-29 | 52 (0.9%) | 38 (0.6%) | 0.30 (-0.05, 0.60) | 28 (0.9%) | 23 (0.9%) | 0.05 (-0.48, 0.54) |
| | 30-39 | 72 (1.5%) | 40 (0.6%) | 0.91 (0.51, 1.31) | 36 (1.2%) | 22 (0.7%) | 0.55 (0.05, 1.05) |
| | 40-49 | 95 (1.4%) | 103 (1.0%) | 0.42 (0.07, 0.77) | 57 (1.3%) | 63 (1.2%) | 0.13 (-0.33, 0.58) |
| | 50-59 | 116 (1.4%) | 200 (1.3%) | 0.04 (-0.25, 0.34) | 94 (1.5%) | 127 (1.5%) | -0.04 (-0.43, 0.38) |
| | 60-69 | 66 (1.3%) | 165 (1.5%) | -0.19 (-0.58, 0.18) | 67 (1.5%) | 143 (1.9%) | -0.31 (-0.75, 0.13) |
| | 70+ | 31 (1.1%) | 78 (1.5%) | -0.34 (-0.78, 0.09) | 30 (1.0%) | 72 (1.3%) | -0.25 (-0.62, 0.18) |

**Table S6: Risk differences of self-reported diagnoses after 6-12 months, comparing SARS-CoV-2 test-positive and test-negative participants, stratified by sex and age group, continued**
(page 2 of 2)

| | | Female (n = 93 494) | | | Male (n = 59 386) | | |
|---|---|---|---|---|---|---|---|
| **Medical diagnosis** | **Age (years)** | **Positive (n, %)** | **Negative (n, %)** | **RD (95% CI)** | **Positive (n, %)** | **Negative (n, %)** | **RD (95% CI)** |
| Chronic fatigue syndrome | 15-19 | 58 (3.9%) | 27 (1.3%) | 2.89 (1.75, 4.12) | 16 (1.9%) | 5 (0.5%) | 1.61 (0.56, 2.90) |
| | 20-29 | 192 (3.3%) | 40 (0.6%) | 2.98 (2.38, 3.51) | 80 (2.6%) | 26 (1.0%) | 1.86 (1.13, 2.58) |
| | 30-39 | 209 (4.3%) | 39 (0.6%) | 4.00 (3.39, 4.66) | 101 (3.4%) | 27 (0.8%) | 2.80 (2.03, 3.57) |
| | 40-49 | 268 (3.9%) | 120 (1.1%) | 2.86 (2.35, 3.43) | 153 (3.6%) | 67 (1.3%) | 2.39 (1.71, 3.03) |
| | 50-59 | 330 (3.9%) | 219 (1.4%) | 2.46 (2.03, 2.93) | 270 (4.3%) | 152 (1.8%) | 2.42 (1.83, 3.04) |
| | 60-69 | 198 (4.0%) | 208 (1.9%) | 1.94 (1.38, 2.54) | 203 (4.6%) | 174 (2.3%) | 2.17 (1.52, 2.77) |
| | 70+ | 169 (6.1%) | 117 (2.3%) | 3.25 (2.40, 4.12) | 154 (5.3%) | 108 (2.0%) | 2.81 (2.05, 3.65) |
| Fibromyalgia | 15-19 | 15 (1.0%) | 6 (0.3%) | 0.75 (0.22, 1.36) | 6 (0.7%) | 5 (0.5%) | 0.24 (-0.54, 0.99) |
| | 20-29 | 36 (0.6%) | 20 (0.3%) | 0.32 (0.06, 0.56) | 20 (0.6%) | 20 (0.7%) | -0.10 (-0.57, 0.36) |
| | 30-39 | 44 (0.9%) | 30 (0.5%) | 0.46 (0.15, 0.80) | 27 (0.9%) | 21 (0.6%) | 0.27 (-0.20, 0.71) |
| | 40-49 | 77 (1.1%) | 92 (0.9%) | 0.25 (-0.06, 0.55) | 49 (1.1%) | 52 (1.0%) | 0.15 (-0.25, 0.55) |
| | 50-59 | 83 (1.0%) | 171 (1.1%) | -0.15 (-0.43, 0.12) | 85 (1.3%) | 114 (1.4%) | -0.02 (-0.41, 0.37) |
| | 60-69 | 52 (1.0%) | 165 (1.5%) | -0.47 (-0.84, -0.11) | 60 (1.4%) | 138 (1.8%) | -0.41 (-0.81, 0.05) |
| | 70+ | 40 (1.4%) | 80 (1.5%) | -0.10 (-0.62, 0.41) | 26 (0.9%) | 72 (1.3%) | -0.38 (-0.78, 0.02) |

**Figure S1: Risk differences of symptoms around the test date, comparing SARS-CoV-2 test-positive and test-negative participants.**

| Symptom | Positive (n,%) | Negative (n,%) | Risk Difference (95% CI) |
|---|---|---|---|
| Dysgeusia | 22,419 (36.8%) | 237 (5.3%) | 34.49 (33.74, 35.28) |
| Dysosmia | 21,884 (35.9%) | 247 (5.5%) | 33.87 (33.06, 34.73) |
| Fever | 33,577 (55.0%) | 1,453 (32.5%) | 23.90 (22.35, 25.28) |
| Chills | 22,800 (37.4%) | 810 (18.1%) | 21.86 (20.59, 23.15) |
| Muscle/joint pain | 25,835 (42.4%) | 1,066 (23.9%) | 21.54 (20.27, 22.99) |
| Reduced appetite | 19,183 (31.4%) | 639 (14.3%) | 19.05 (17.94, 20.13) |
| Reduced strength legs/arms | 15,770 (25.9%) | 431 (9.7%) | 17.33 (16.30, 18.28) |
| Hot flushes/sweat | 21,220 (34.8%) | 904 (20.2%) | 16.37 (14.87, 17.73) |
| Fatigue/exhaustion | 28,779 (47.2%) | 1,515 (33.9%) | 14.39 (12.85, 16.00) |
| Dizziness | 13,440 (22.0%) | 452 (10.1%) | 13.77 (12.78, 14.73) |
| Dyspnea | 12,213 (20.0%) | 370 (8.3%) | 13.60 (12.62, 14.44) |
| Headache | 26,880 (44.1%) | 1,481 (33.2%) | 11.59 (10.00, 13.16) |
| Chest pain | 9,003 (14.8%) | 279 (6.2%) | 10.01 (9.19, 10.73) |
| Nausea | 7,450 (12.2%) | 320 (7.2%) | 6.41 (5.67, 7.25) |
| Sleeping legs/arms | 5,524 (9.1%) | 162 (3.6%) | 6.19 (5.59, 6.75) |
| Diarrhoea | 6,559 (10.8%) | 301 (6.7%) | 4.93 (4.17, 5.65) |
| Abdominal pain | 5,103 (8.4%) | 279 (6.2%) | 3.17 (2.42, 3.87) |
| Red runny eyes | 4,126 (6.8%) | 208 (4.7%) | 2.70 (2.01, 3.35) |
| Cough | 21,917 (35.9%) | 1,772 (39.7%) | −1.79 (−3.39, −0.27) |
| Runny nose | 15,654 (25.7%) | 1,770 (39.6%) | −11.98 (−13.58, −10.22) |
| Sore throat | 17,902 (29.3%) | 2,183 (48.9%) | −16.93 (−18.51, −15.27) |

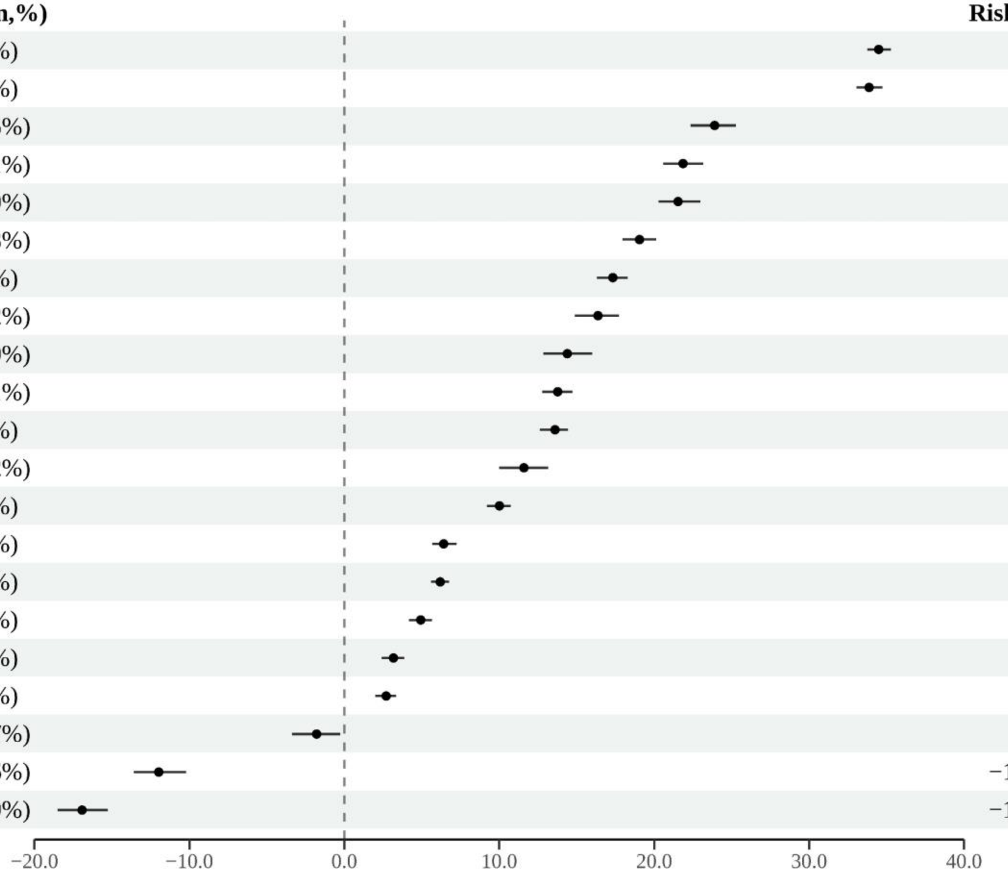

Note: "Around the test date" is defined as from one week prior to the test date and until four weeks after. For symptoms around the test date, test-negatives with symptoms as indication for testing are used as control population. Risk differences with 95% confidence intervals were adjusted for age, sex, comorbidities, obesity, healthcare-occupation and time after testing (in months).

**Figure S2: Risk differences of symptoms 6-12 months after test, comparing hospitalized and non-hospitalized SARS-CoV-2 test-positive participants.**

| Symptom | Hospitalized (n,%) | Non−hospitalized (n,%) | Risk Difference (95% CI) |
|---|---|---|---|
| Fatigue/exhaustion | 397 (16.4%) | 6,402 (10.9%) | 8.64 (6.70, 10.74) |
| Reduced strength legs/arms | 302 (12.5%) | 3,079 (5.3%) | 7.13 (5.55, 8.66) |
| Dyspnea | 274 (11.3%) | 3,003 (5.1%) | 6.71 (5.17, 8.39) |
| Muscle/joint pain | 201 (8.3%) | 3,016 (5.1%) | 5.05 (3.57, 6.72) |
| Headache | 180 (7.4%) | 3,560 (6.1%) | 4.58 (2.86, 6.31) |
| Sleeping legs/arms | 203 (8.4%) | 2,638 (4.5%) | 4.25 (2.87, 5.68) |
| Dizziness | 158 (6.5%) | 2,272 (3.9%) | 4.11 (2.64, 5.72) |
| Chest pain | 121 (5.0%) | 1,574 (2.7%) | 4.02 (2.69, 5.38) |
| Cough | 151 (6.2%) | 2,805 (4.8%) | 2.81 (1.45, 4.36) |
| Hot flushes/sweat | 108 (4.5%) | 1,939 (3.3%) | 2.53 (1.34, 3.86) |
| Nausea | 69 (2.9%) | 1,110 (1.9%) | 2.38 (1.24, 3.53) |
| Abdominal pain | 71 (2.9%) | 1,170 (2.0%) | 1.88 (0.93, 2.87) |
| Reduced appetite | 90 (3.7%) | 1,682 (2.9%) | 1.78 (0.63, 2.95) |
| Sore throat | 95 (3.9%) | 2,187 (3.7%) | 1.52 (0.15, 2.80) |
| Fever | 68 (2.8%) | 1,294 (2.2%) | 1.44 (0.43, 2.46) |
| Red runny eyes | 58 (2.4%) | 764 (1.3%) | 1.41 (0.62, 2.32) |
| Chills | 57 (2.4%) | 909 (1.6%) | 1.38 (0.53, 2.28) |
| Runny nose | 92 (3.8%) | 2,284 (3.9%) | 1.15 (−0.12, 2.36) |
| Diarrhoea | 57 (2.4%) | 1,065 (1.8%) | 0.98 (0.16, 1.79) |
| Dysgeusia | 184 (7.6%) | 5,181 (8.8%) | 0.97 (−0.65, 2.69) |
| Dysosmia | 188 (7.8%) | 6,486 (11.1%) | −0.52 (−2.27, 1.21) |

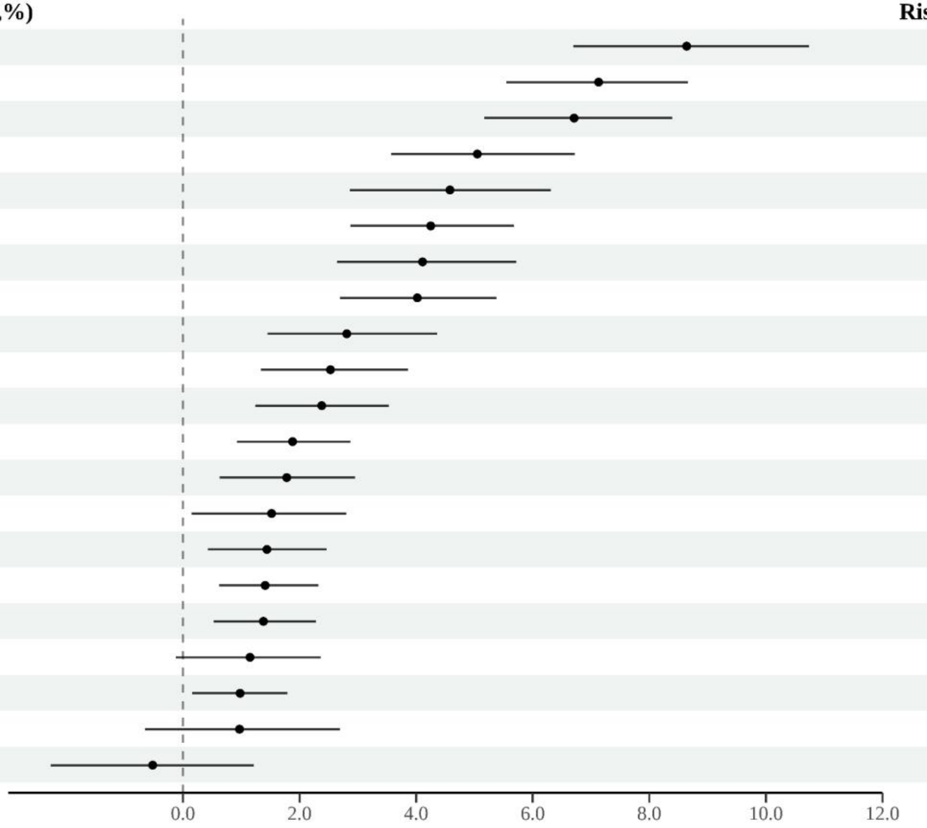

**Figure S3: Risk differences self-reported health problems with new onset from the test date and up to after 6-12 months, comparing SARS-CoV-2 test-positive and test-negative participants, stratified by sex and age group**

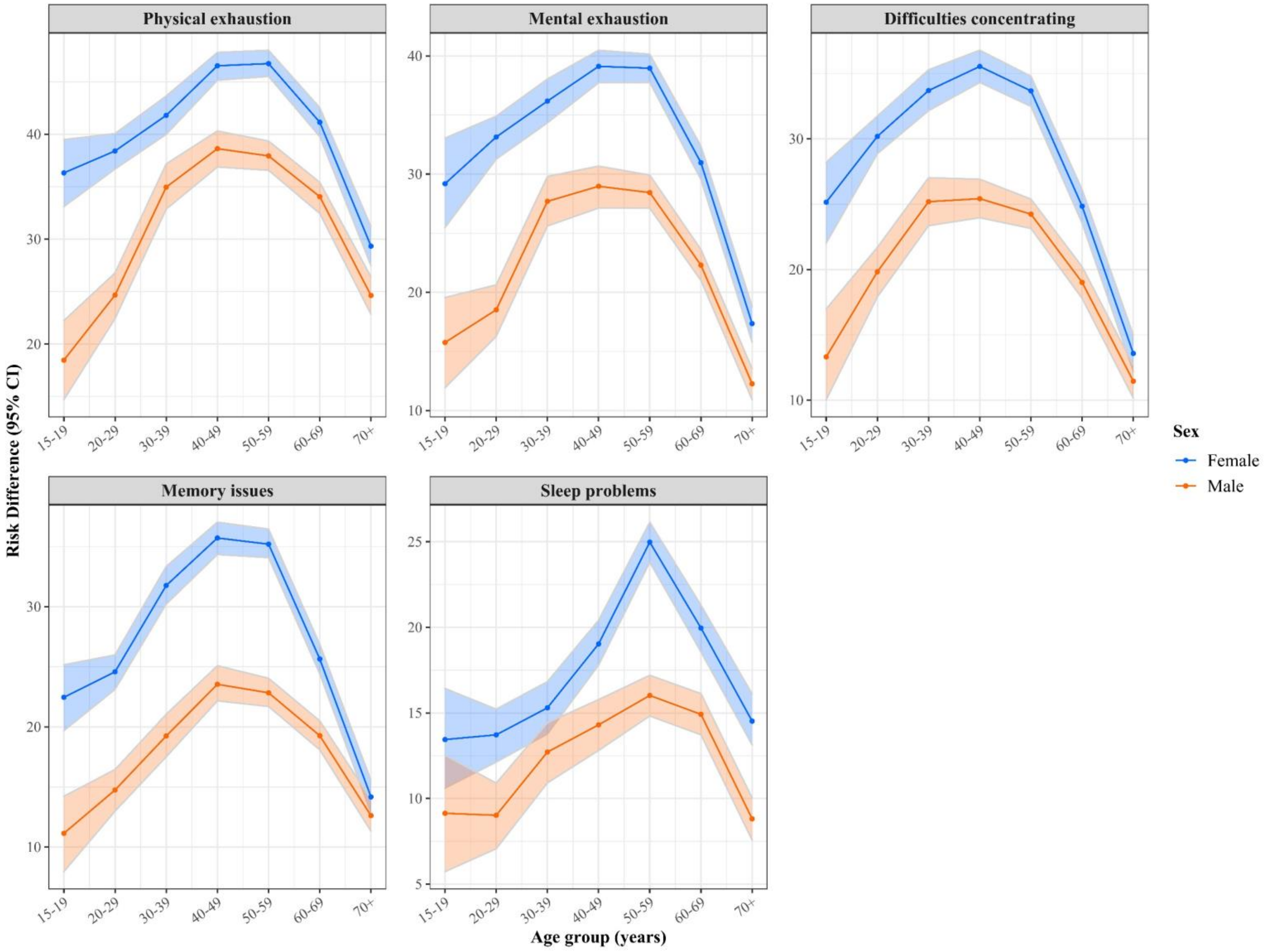

**Figure S4: Risk differences self-reported diagnoses with new onset from the test date and up to after 6-12 months, comparing SARS-CoV-2 test-positive and test-negative participants, stratified by sex and age group**

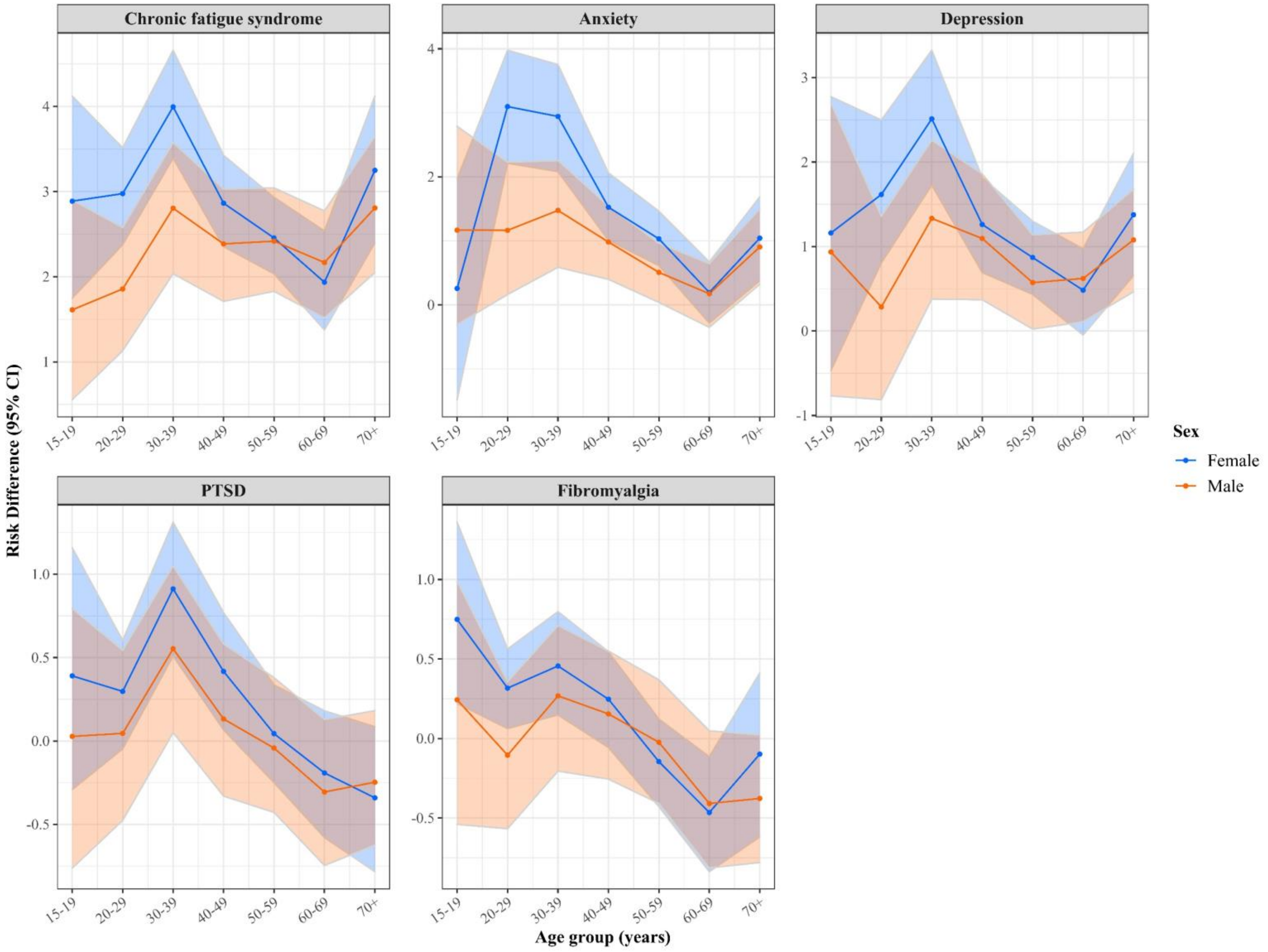
